# The Gut Microbiome and their Metabolites in Human Blood Pressure Variability

**DOI:** 10.1101/2022.03.15.22272376

**Authors:** Evany Dinakis, Michael Nakai, Paul Gill, Rosilene Ribeiro, Stephanie Yiallourou, Yusuke Sata, Jane Muir, Melinda Carrington, Geoffrey A. Head, David M. Kaye, Francine Z. Marques

## Abstract

Blood pressure (BP) variability is an independent risk factor for cardiovascular events. Recent evidence supports a role for the gut microbiota in BP regulation. However, whether the gut microbiome is associated with BP variability is yet to be determined. Here, we aimed to investigate the interplay between the gut microbiome and their metabolites in relation to BP variability. Ambulatory BP monitoring was performed in 69 participants from Australia (55.1% women; mean±SD 59.8±7.26-years old, 25.2±2.83 kg/m^2^). This data was used to determine night-time dipping, morning BP surge (MBPS) and BP variability as standard deviation (SD). The gut microbiome was determined by 16S rRNA sequencing, and metabolite levels by gas chromatography. We identified specific taxa associated with systolic BP variability, night-time dipping and MBPS. Notably, *Alistipesfinegoldii* and *Lactobacillus spp*. were only present in participants within the normal ranges of BP variability, MBPS and dipping, while *Prevotella spp*. and various *Clostridium* spp. were found to be present in extreme dippers and the highest quartiles of BP SD and MBPS. There was a negative association between MBPS and microbial α-diversity (r=-0.244, *P*=0.046). MBPS was also negatively associated with total levels of microbial metabolites called short-chain fatty acids (SCFAs) in the plasma (r=-0.305, *P*=0.020), particularly acetate (r=-0.311, *P*=0.017). In conclusion, gut microbiome diversity, levels of microbial metabolites, and the bacteria *Alistipesfinegoldii* and *Lactobacillus* were associated with lower BP variability, and *Clostridium* and *Prevotella* with higher BP variability. Thus, our findings suggest the gut microbiome and metabolites may be involved in the regulation of BP variability.

## Introduction

Over a 24-hour period, blood pressure (BP) follows a natural pattern of diurnal variation, independent of internal and external stimuli.^1^ Typically, BP oscillates regularly through the day and night cycle. BP undergoes a prominent decline at night during the sleep cycle, referred to as ‘night-time dipping’, followed by an abrupt, steep acceleration in the morning following rising, known as ‘morning surge’.^2,3^ Night-time dipping in systolic BP, defined as the difference between the mean systolic pressure of day and night BP, is recognised as a measure of cardiovascular risk,^4^ with an accepted normal range between 10-20%.^3,5^ Thus, individuals who do not display this pattern of dipping (non-dippers), or instead surpass the acceptable dipping range (extreme dippers) have a greater risk of mortality and morbidity.^6,7^ Similarly, an exaggerated morning surge in systolic BP has also been identified as a risk factor in the development of cardiovascular disease (CVD), particularly in hypertensive patients.^8,9^

The gut microbiota is the community of microorganisms that resides in the intestine.^10,11^ Over the past decade, the gut microbiota has emerged as a key player in BP regulation.^11^ Emerging data demonstrate that the human gut microbiome, similar to 24-hour BP, exhibits diurnal variations^12,13^ and is tightly synchronized to the host circadian rhythm.^14^ However, circadian disruption, whether central (i.e. light-dark shift, artificial light at night) or peripheral (i.e. timing of food consumption) in nature, can lead to imbalances in intestinal microbiota, gut dysbiosis and changes in biological components of the microbiota themselves.^14^ In turn, this may influence both metabolic and inflammatory pathways that alter the natural pattern of BP variability. ^13^ Such alterations, primarily gut dysbiosis, have previously been associated with elevated BP in several human studies.^10,11,15^ Experiments using faecal microbiota transplantation to germ-free animals have demonstrated the gut microbiota is not merely associated with but can in fact increase BP.^16,17^ This may be attributed to mechanisms involving gut microbial-derived metabolites, such as short-chain fatty acids (SCFAs), which are produced during fermentation of dietary fibre by intestinal bacteria.^11^ The production of these bacterial metabolites is also rhythmic,^18^ which is critical as recent studies suggest that the host’s diurnal rhythms may be driven by gut microbiota and their metabolites.^18,19^ However, whether the gut microbiota and their metabolites are associated with changes in the natural circadian pattern of BP regulation, particularly in humans, remains to be elucidated. Here, we aimed to study this phenomenon in normotensive and untreated hypertensive participants with well-characterised BP and the gut microbiome, their metabolites and receptors.

## Methods

### Participants and recruitment

The cohort was recently described elsewhere.^20^ Briefly, a total of 76 participants were recruited from metropolitan (n=41 Baker Institute and Alfred Hospital, Melbourne) and regional (n=35, Shepparton, Victoria) areas between October-2016 and April-2018 for this observational study. Inclusion criteria were defined as: aged 40-70 years, either sex, body mass index (BMI) 18.5-30 kg/m^2^, and not using BP-lowering medication. Exclusion criteria included gastrointestinal disease (including history of intestinal surgery, inflammatory bowel disease, celiac disease, lactose intolerance, chronic pancreatitis or other malabsorption disorder), diabetes (type 1 and 2), chronic kidney disease, and probiotic or antibiotic use in the past 3 months. Two participants were excluded due to high BMI and four were excluded due to incomplete 24-hour BP measurements. Following exclusions, a total of 70 participants remained: 40 in the metropolitan clinics and 30 in the regional clinic. This study complied with the Declaration of Helsinki, and was approved by the human research ethics committee of the Alfred Hospital (approval 415/16). All participants provided informed consent. The study was registered in the Australian New Zealand Clinical Trials Registry under ACTRN12620000958987.

### BP measurement and its variability calculation

Participants were fitted with a calibrated ambulatory BP monitoring device (AND or SpaceLabs) for 24-hours. Data from two participants for BP variability, one participant for night-time dipping and three participants for morning BP surge (MBPS) data were excluded due to lack of specific BP timepoints required to calculate these metrics. BP variability is represented as the standard deviation (SD) of total day systolic (divided into quartiles: Q1: <9.76; Q2: 9.76–11.32; Q3: 11.32-14.14; Q4: >14.14), night systolic (Q1: <9.75; Q2: 9.75– 11.38; Q3: 11.38-13.65; Q4: >13.65), and overall fitted data (Q1: <11.94; Q2: 11.94–13.86; Q3: 13.86-16.30; Q4: >16.30).

Night-time dipping and morning surge data were manually calculated as previously described.^21^ Briefly, sleep BP was defined as the BP average from the time when the participant went to bed until rising (11:00pm – 6:00am) and awake BP as the BP average of the remainder of the day (7:00am – 10:00pm). Participants were then classified according to the percentage of night-time dipping [100x (1-sleep BP/awake BP)] as follows: extreme dippers (night-time dip ≥20%), dippers (night-time dip ≥10% but <20%), or non-dippers (night-time dip ≥0% but <10%).

Morning BP was defined as the BP average during the first two hours after rising (7:00am – 9:00am; 5 readings); preawake BP was defined as the average BP during the two hours prior to rising (4:00am – 6:00am; 5 readings); the lowest BP was defined as the average BP of the lowest night-time reading, and the reading immediately before and after (3 readings). Sleep-through MBPS was calculated as: morning BP – lowest BP, and prewaking MBPS as: morning BP – preawake BP. The cohort was then divided into quartiles for each MBPS parameter: sleep-through MBPS (quartiles: Q1: <9.40; Q2: 9.40–17.17; Q3: 17.17-23.45; Q4: >23.45) and prewaking MBPS (Q1: <3.93; Q2: 3.93–12.73; Q3: 12.73-18.33; Q4: >18.33).

### Food Frequency Questionnaire

Dietary intake over a 12-month period was assessed using the Dietary Questionnaire for Epidemiological Studies version 3.2, a self-administered questionnaire developed by the Cancer Council Victoria that allows for a reflection of dietary intake of the Australian population. ^22^ Subtypes of dietary fibre (insoluble and soluble fibre, resistant starch) were estimated using previously described tools.^23^ The quality of participants’ diet was also measured as Australian Dietary Guideline Index (DGI-2013)^24,25^ as described previously.^20,26^

### Faecal DNA extraction, library preparation and sequencing

This study followed guidelines for gut microbiota studies in hypertension^27^ and the Strengthening the Organization and Reporting of Microbiome Studies (STORMS) reporting^28^ (checklist available at ^20^). Sample collection, DNA extraction, library preparation and sequencing were described in detail previously.^20^ Briefly, DNA from the stool samples was extracted using the DNeasy PowerSoil DNA isolation kit (Qiagen). The V4-V5 region of the bacterial 16S rRNA was amplified by PCR using the Earth Microbiome Project^29^ protocol. The libraries were sequenced in an Illumina MiSeq sequencer (300 bp paired-end reads). To increase the reproducibility of the findings, all samples were independently sequenced twice. These technical duplicated samples were combined for the analyses described below.

### Bioinformatic analyses of gut microbiome

Anonymized microbiome data and materials have been made publicly available at the NCBI Sequence Read Archive (SRA) database under access PRJNA722359.^20^ Sequence reads from samples were first analysed using the QIIME2 framework.^30^ Samples were rarefied at 29,000 reads per sample. β diversity metrics (showing differences in the composition of the microbiome) were generated from the rarefied samples, including unweighted and weighted Unifrac metrics as well as associated Principal Coordinate Analysis (PCoA) distance tables. Taxonomic assignment used a nai[ve Bayes classifier to label ASVs (via q2-feature-classifier^31^), trained against the SILVA database (version 138) 99% OTU reference sequences specific for bacterial V4-V5 rRNA regions. Further analyses were performed on MicrobiomeAnalyst from the rarefied samples, including α diversity (showing differences in the number of taxa and/or how evenly distributed they were) and abundance profiling. Features with a minimum of four counts occurring at a prevalence of 10% of samples were included. Data was scaled using the Total Sum Scaling (TSS) normalisation method.^32^ Differential taxa analysis was performed using edgeR (adjusted *P*-value cut-off <0.05 on feature-level) on MicrobiomeAnalyst.^33,34^ One participant was excluded from all analyses due to low total number of sequencing reads (<10,000), thus resulting in 69 participants that were included the analyses.

### Short-chain fatty acids measurement

Briefly, plasma SCFAs were measured in 200 µL and faecal SCFAs were measured from 1 g of faecal sample, all in triplicates, as previously published,^20^ in an Agilent GC6890 coupled to a flame-ionisation detector.^35,36^ A coefficient of variation of <10% within triplicate samples was used as a quality control measure.

### Blood expression of SCFA receptors and transporters

Using knockout mice, we have previously shown that all three main SCFA-sensing receptors GPR41 (*FFAR3)*, GPR43 (*FFAR2*) and GPR109A (*HCAR2*) have a role in cardiovascular dysfunction.^10^ As these receptors are highly expressed in immune cells,^37^ we quantified the expression of the mRNA of the three receptors in circulating immune cells in 50 participants. TaqMan assays were used in a QuantStudio 6 Flex Real-Time PCR (qPCR) system (all ThermoFisher Scientific), with glyceraldehyde 3-phosphate dehydrogenase (*GAPDH*) and β-actin (*ACTB*) as housekeeping genes. All expression experiments were run in duplicates and significance was assessed by 2^-ΔΔ*C*^T method.

### Statistical analyses

Microbiome data was analysed as explained above. Data were analysed blind. GraphPad Prism (version 8) package was used for graphing, and SPSS for Windows (release 25) for statistical analyses. Non-parametric tests were used in the case of non-normally distributed data. A one-way ANOVA was performed on quartile and night-time dipping data, while a two-tail independent sample t-test was used to compare Q1-Q3 to Q4 for each parameter. Normally distributed α diversity score correlations were performed using Pearson’s correlation coefficient, whereas non-normally distributed correlation data were performed using Spearman’s correlation coefficient. Further analyses were conducted using step-wise multiple linear regression models for acetate, butyrate, propionate, *FFAR3, FFAR2* and *HCAR2* levels. These models had clinical (age, sex, body mass index) variables as independent parameters (criteria of F-entry probability: 0.15, removal: 0.20) in SPSS for Windows (release 25). Data are presented as mean ± SD unless otherwise specified, and those with a *P*<0.05 were considered significant.

## Results

### Baseline characteristics

Tables 1 and 2 summarise the baseline characteristics of the participants for the SD, night-time dipping and MBPS data respectively, across quartiles and night-time dipping status. Besides the difference between quartiles for each of the BP variability parameters studied, only sex was different across day and night SD quartiles, with less women in Q4 group. A total of over 4.3 million sequencing reads were denoised, merged and underwent chimera filtering, resulting in an average read count of 63,000 per sample. Samples were rarefied to 29,000 reads to allow for consistent and plateauing diversity metrics (Online Supplementary Figure S1).

**Table 1.** Demographics and clinical characteristics of participants for standard deviation (SD) and morning blood pressure surge (MBPS).

| Variable | Quartile 1 | Quartile 2 | Quartile 3 | Quartile 4 | P-value |
| --- | --- | --- | --- | --- | --- |
| *SD of day data fitted* | **<9.76** | **9.76–11.32** | **11.32-14.14** | **>14.14** | **<0.001** |
| Age (y) | 56.4±6.54 | 58.0±8.36 | 62.1±7.14 | 61.4±6.14 | 0.479 |
| Sex (% Female) | 62.5% | 70.6% | 38.9% | 47.1% | **0.02** |
| BMI | 24.9±2.75 | 25.8±2.49 | 25.0±3.02 | 24.9±3.24 | 0.623 |
| *SD of night data fitted* | **<9.75** | **9.75–11.38** | **11.38-13.65** | **>13.65** | **<0.001** |
| Age (y) | 60.1±7.56 | 56.5±8.67 | 60.4±7.02 | 61.2±5.49 | 0.254 |
| Sex (% Female) | 68.8% | 82.4% | 50% | 17.6% | **<0.001** |
| BMI | 24.2±2.93 | 24.9±2.28 | 25.8±3.16 | 25.6±2.92 | 0.382 |
| *SD of total data fitted* | **<11.94** | **11.94-13.86** | **13.86-16.30** | **>16.30** | **<0.001** |
| Age (y) | 57.6±6.70 | 61.5±8.25 | 59.1±7.59 | 59.9±6.71 | 0.073 |
| Sex (% Female) | 81.3% | 47.1% | 61.1% | 29.4% | 0.236 |
| BMI | 25.5±2.95 | 25.5±2.86 | 24.4±2.71 | 25.2±3.00 | 0.800 |
| *Sleep-through MBPS (range)* | **2.30±5.67** | **12.8±2.39** | **17.17-23.45** | **>23.45** | **<0.001** |
|  | **(<9.40)** | **(9.40–17.17)** | **(19.9±1.78)** | **(36.8±9.63)** |  |
| Age (y) | 61.4±7.52 | 57.0±8.02 | 60.6±7.11 | 58.6±6.34 | 0.307 |
| Sex (% Female) | 50% | 62.5% | 61.1% | 41.2% | 0.577 |
| BMI | 26.0±3.13 | 25.2±2.72 | 24.6±2.46 | 25.0±3.22 | 0.556 |
| *Pre-waking MBPS (range)* | **-3.10±7.53**<br>**(<3.93)** | **8.14±2.48**<br>**(3.93–12.73)** | **15.7±1.58**<br>**(12.73-18.33)** | **27.7±7.48**<br>**(>18.33)** | **<0.001** |
| Age (y) | 61.6±6.32 | 58.9±9.65 | 56.4±6.54 | 60.5±6.26 | 0.165 |
| Sex (% Female) | 36.8% | 71.4% | 58.8% | 52.9% | 0.258 |
| BMI | 25.6±3.25 | 25.2±2.96 | 24.8±2.24 | 25.0±3.09 | 0.853 |

Data are shown as mean±standard deviation or numbers and percentages. Legend: standard deviation, SD; body mass index, BMI; systolic blood pressure, SBP. *P*-value from one-way ANOVA, in bold *P*<0.05.

**Table 2.** Demographics and clinical characteristics of participants for night-time dipping data.

| Variable | Extreme Dippers | Dippers | Non-Dippers | *P*-value |
| --- | --- | --- | --- | --- |
| Sample size (n, %) | 7 (10.1%) | 43 (62.3%) | 19 (27.5%) |  |
| Age (y) | 57.1±5.08 | 58.7±7.69 | 62.5±6.38 | 0.166 |
| Sex (% Female) | 57.1% | 53.5% | 57.9% | 0.686 |
| BMI | 23.9±2.91 | 25.3±2.59 | 25.3±3.33 | 0.231 |
| Night-time dipping (%) | 23.0±3.83 | 15.1±2.73 | 4.88±3.45 | **<0.001** |

Data are shown as mean±standard deviation or numbers and percentages. Legend: body mass index, BMI; systolic blood pressure, SBP. *P*-value from one-way ANOVA, in bold *P*<0.05.

### Dietary Food Intake

There was no difference in total fibre, insoluble and soluble fibre, as well as resistant starches between all three metrics of SD BP data, night-time dipping and both sleep-through and prewaking MBPS (data not shown, all *P*>0.05). However, we identified a negative correlation between sleep-through MBPS and the Australian Dietary Score (r=-0.29, *P*=0.017), which remained significant after adjustment for age, BMI and sex (β=-0.522, *P*=0.017). All other comparisons with BP variability metrics were not significant.

### BP variability and the gut microbiome

We found no association between three metrics of α-diversity: observed OTUs, Chao1 (measure of species richness) and Shannon index (measure of taxonomic richness and evenness) and BP variability, including SD of day, night and total data fitted (Figures 1A, B, C, Figure S2). These findings were validated in correlations between α-diversity and all three datasets of BP variability as continuous variables (Online Supplementary Table S1). We obtained similar results when assessing β-diversity; both unweighted and weighted UniFrac distances showed no significant clustering patterns between Q1-Q3 versus Q4 of all three datasets of BP variability (Figure S3).

**Figure 1.**
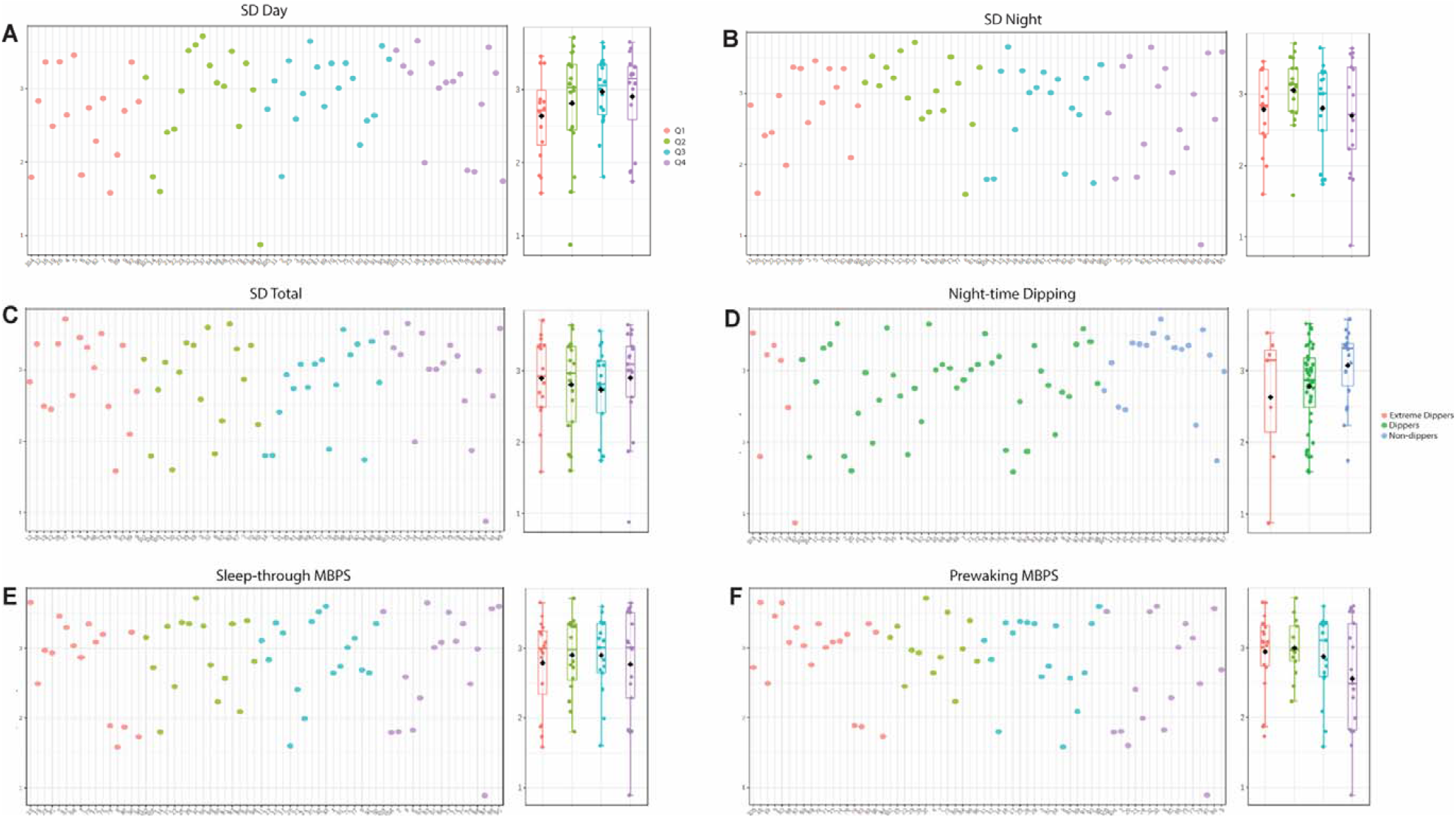
α-diversity profiles and box plots of blood pressure (BP) variability. α-diversity profiling showing Shannon index, which combines both richness and evenness, for A) BP standard deviation (SD) day, B) night and C) total, D) night-time dipping, E) sleep-through and F) prewaking morning blood pressure surge (MBPS). Box plot data presented as median and inter-quartile range (IQR). All *P*>0.05.

We identified 46 unique microbial taxa that were more prevalent in the lowest quartiles of BP SD (Q1-Q3) versus the highest quartile (Q4) (Table S2, main findings summarised in Figure 2). We found that *Alistipesfinegoldii* (Figure 2A and B respectively, *P*=0.032, *P*=0.018), *Lactobacillus* spp. (Figure 2C, *P*=0.025), uncultured *Acetivibrio* spp. (Figures 2D-E, *P*=0.033, *P*=0.0021) and *Azospirillum* spp. (Figure 2F, *P*=0.0013) were more abundant in the lowest quartiles of SD data compared to Q4. Conversely, we found that participants in the highest total SD quartile had higher abundance levels of *Clostridium* spp. including *ClostridialesvadinBB60group* and uncultured *Clostridiumsp*_*6* (Figures 2G and H respectively, *P*<0.001, *P*=0.0056).

**Figure 2.**
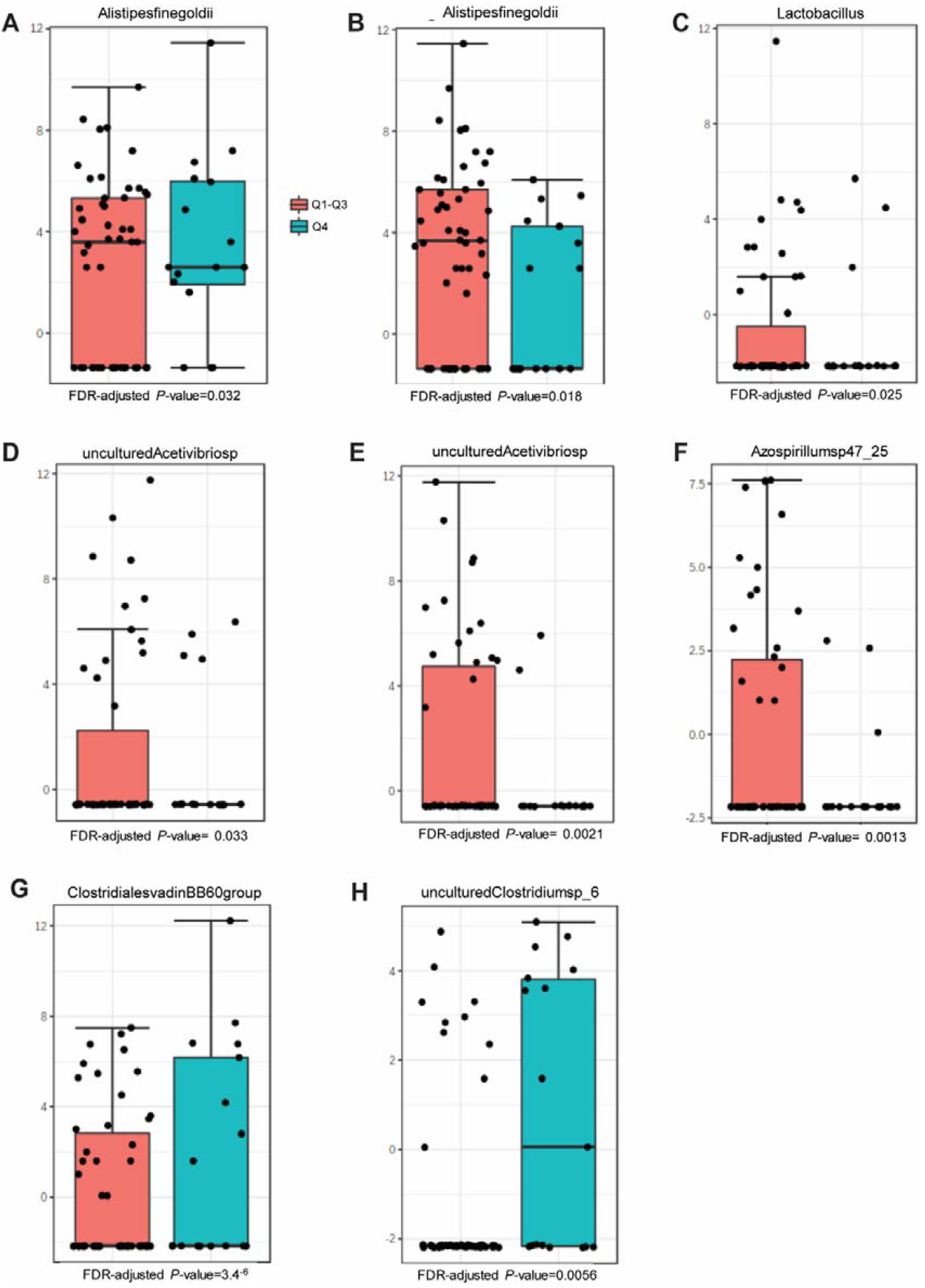
Differential abundance analysis of specific bacterial taxa for blood pressure standard deviation (SD). Differentially abundant bacterial taxa in Q1-Q3 compared to Q4 for A) SD of day data fitted, B, C, D) SD of night data fitted and E, F) SD of total data fitted. Differentially abundant bacterial taxa in Q4 compared to G, H) Q1-Q3 for SD of total data fitted. The data presented is presented as log-transformed counts. See Table S2 for more details. False discovery rate adjusted *P*-value cut-off=0.05. Box plot data presented as median and IQR.

### Night-time dipping and the gut microbiome

There was no association between α-diversity and night-time dipping both as a categorical (Figure 1D and S4) and continuous variable (Table S1). We also did not identify any significant clustering patterns between extreme dippers, dippers and non-dippers both in unweighted and weighted PCoA plots (Figure S5). However, we still identified taxonomic differences between normal and irregular dipping patterns (Tables S3-S5). Compared to extreme dippers, it was found that dippers had differentially abundant levels of *Gaboniamassiliensis* (Figure 3A, *P*=0.041). However, extreme dippers exhibited higher levels of various *Clostridium* spp., specifically *ClostridialesvadinBB60group* (Figure 3B, *P*<0.001), as well as higher levels of *Prevotella* spp. including *PrevotellacaeNK3B31group* (Figure 3C, *P*=0.018). When comparing dippers to non-dippers, we found that *Lactobacillus* spp. were more abundant in dippers (Figure 3D, *P*<0.001), however *Clostridium* spp. including uncultured *Clostridiumsp_4* and *Ruminiclostridium6* were higher in participants with a non-dipping profile (Figures 3E and Frespectively, *P*<0.001, *P*=0.046). When comparing all three night-time dipping profiles, dippers had differentially abundant levels of *Lactobacillus* spp. compared to both extreme- and non-dippers (Figure 3G, *P*=0.030), whereas extreme dippers had significantly more abundant levels of *PrevotellacaeNK3B31group* spp. compared to dippers and non-dippers (Figure 3H, *P*=0.017). Despite all three dipping profiles exhibiting levels of *ClostridialesvadinBB60group* spp., it was extreme dippers who had the greatest levels compared to dippers and non-dippers (Figure 3I, *P*<0.001).

**Figure 3.**
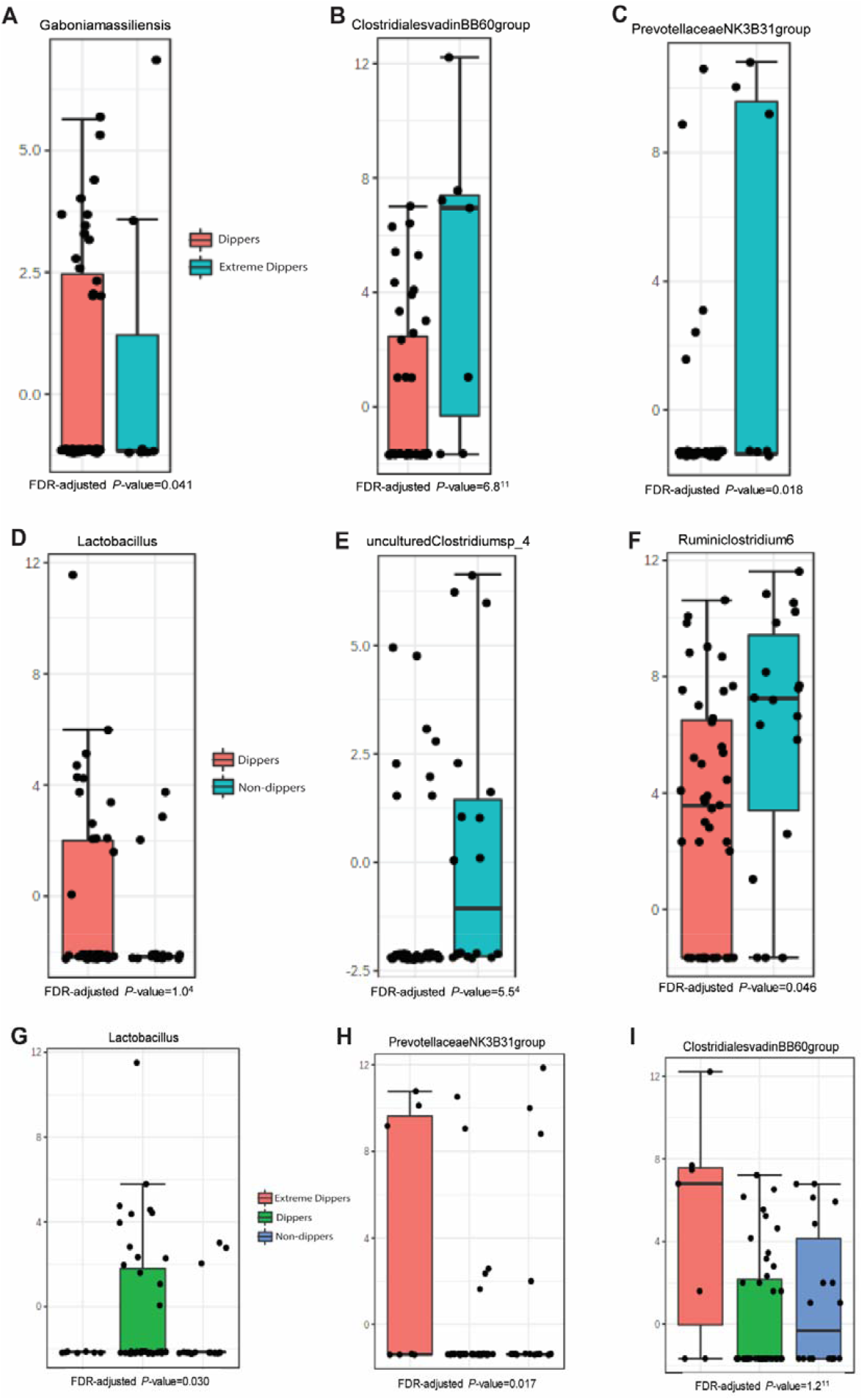
Differential abundance analysis of specific bacterial taxa for night-time dipping. Differentially abundant bacterial taxa in A, B, C) dippers compared to extreme dippers, D, E, F) dippers compared to non-dippers and G, H, I) all classifications of dipping. The data is presented as log-transformed counts. See Tables S3-S5 for more details. False discovery rate adjusted *P*-value cut-off=0.05. Box plot data presented as median and IQR.

### MBPS and the gut microbiome

α-diversity profiling analyses indicated that, similarly to both BP SD and night-time dipping data, no significant associations were reported between metrics of α-diversity and MBPS quartiles (Figure 1E, F and S6). Nevertheless, as a continuous variable, prewaking MBPS was negatively correlated with Shannon Index (Table S1, r=-0.24, *P*=0.046), which remained significant after performing step-wise regression analyses adjusted by age, sex and BMI (β=-6.769, P=0.017). We identified no significant clustering patterns in both β-diversity metrics of unweighted and weighted UniFrac distances for sleep-through and prewaking MBPS between Q1-3 and Q4 (Figure S7). However, some specific taxa were more prevalent in the three lowest quartiles or the highest quartile of both sleep-through and prewaking MBPS (Figure 4 and Table S6). Participants who were classified as having sleep-through MBPS in the three lowest quartiles had higher levels of uncultured *Acetivibriosp* (Figure 4A, *P*=0.0089), as well as a specific taxa *Alistipesfinegoldii* (Figure 4B, *P*=0.029), whereas participants in the highest quartile of sleep-through MBPS had higher levels of *Clostridium* spp., specifically *ClostridialesvadinBB60group* (Figure 4C, *P*<0.001). Moreover, participants in the three lowest quartiles of prewaking MBPS had higher levels of *Lactobacillus* (Figure 4D, *P*<0.001). Participants in the highest quartile of prewaking MBPS however, exhibited higher levels of *Clostridium* spp. and *Prevotella9* (Figure 4E and F respectively, *P*<0.001, *P*=0.0066).

**Figure 4.**
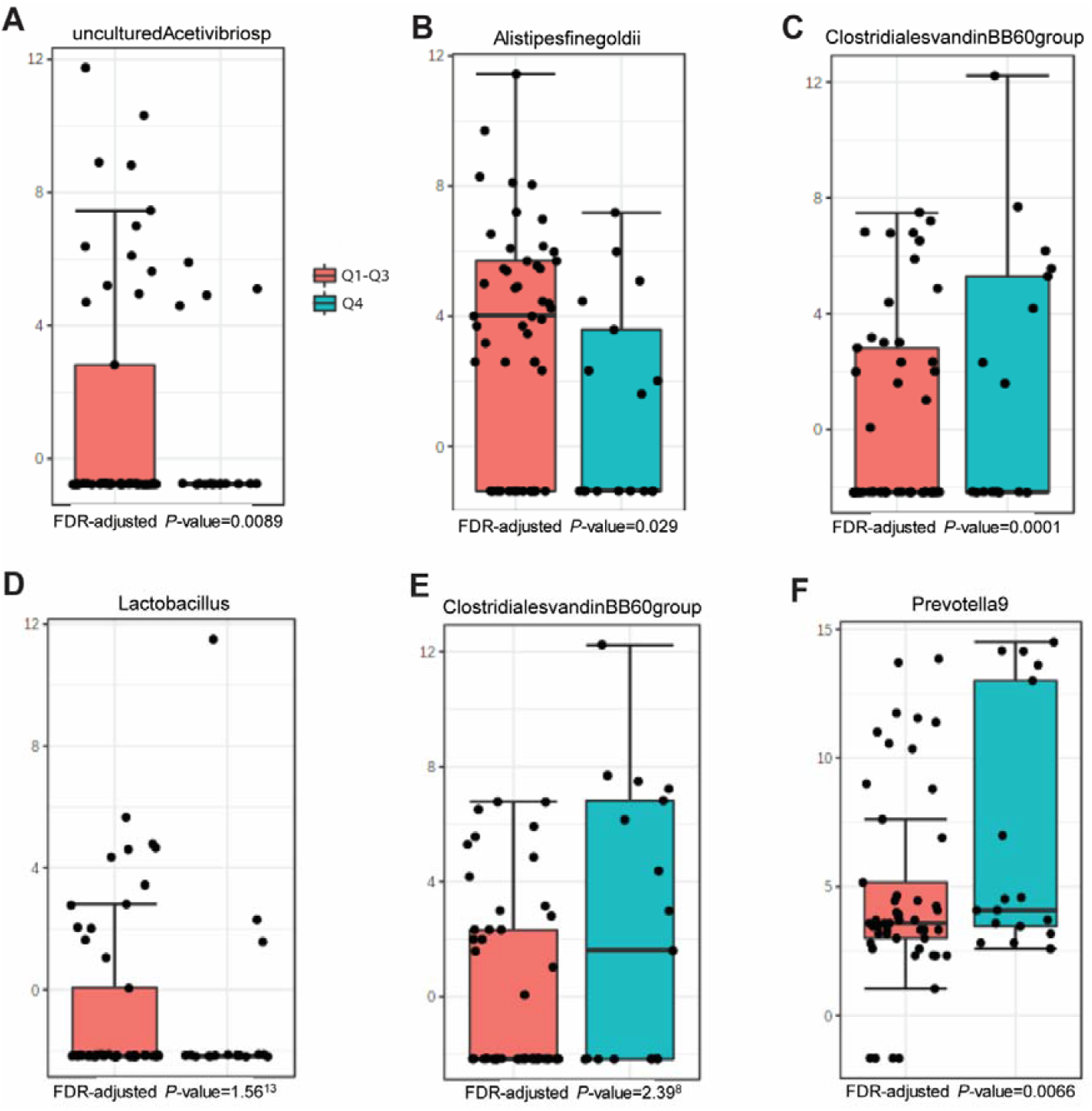
Differential abundance analysis of specific bacterial taxa for sleep-through and prewaking morning blood pressure surge (MBPS). Differentially abundant bacterial taxa in Q1-Q3 compared to Q4. The data presented is presented as log-transformed counts. See Table S6 for more details. False discovery rate adjusted *P*-value cut-off=0.05. Box plot data presented as median and IQR.

### Short-chain fatty acids and receptors

We then quantified the levels of both faecal and circulatory SCFAs in relation to BP SD, night-time dipping and MBPS (Table S7). We identified a negative correlation between plasma total SCFAs and prewaking MBPS (r=-0.31, *P*=0.020; regression analyses β=-0.066, *P*=0.02), driven by plasma acetate levels (r=-0.31, *P*=0.017; regression analyses β=-0.072, *P*=0.017). We then analysed the expression of the SCFA receptors GPR41 (*FFAR3*), GPR43 (*FFAR2*) and GPR109A (*HCAR2*) in circulating immune cells in relation to BP SD, night-time dipping and MBPS. We identified a negative correlation between all three metrics of SD data and *FFAR2* mRNA (Table S8, Figure S8A-C), indicating that participants with higher BP SD had overall lower expression levels of *FFAR2*. Similarly, we found that participants with a greater percentage in night-time dipping had higher levels of *FFAR2* (P=0.0087, Table S8, Figure S8D).

## Discussion

Through a combination of gut microbiome sequencing, metabolite and receptor quantification, and 24-hour ambulatory blood pressure monitoring, we were able to uncover novel relationships with human BP variability represented here as SD, night-time dipping and morning surge. In particular, we discovered certain microbial taxa were associated with BP variability, such as *Lactobacillus, Alistipesfinegoldii, Clostridium* and *Prevotella* spp., as well as associations with microbial diversity and metabolites, particularly the SCFA acetate and its receptor GPR43 (summarised in Figure S9).

A growing body of evidence suggests that dysbiosis of the gut microbiota has a fundamental role in the development of CVD.^38^ Importantly, recent studies support a causal relationship between the gut microbiota and both experimental and human hypertension.^10,11^ Thus, coupled with tight circadian synchronization,^14^ a link between the gut microbiota and diurnal variations in 24-hour BP would be expected. Yet, there is limited evidence implicating the gut microbiota in the host’s circadian rhythms.^18^ In this study, we found no association between microbial α-diversity and BP variability (SD) and night-time dipping. However, we identified a negative association between Shannon Index and MBPS. Differently from Chao1 and observed OTUs, which are both metrics of richness, Shannon Index considers both microbial richness and evenness. Thus, this may suggest that lower diversity and reduced distribution of microbes are present in faecal samples from those with exaggerated MBPS. This is in agreement with a recent cross-sectional study which analysed the human gut microbiome and a multitude of host factors including BP, which was also negatively correlated with Shannon diversity.^39^ Diet quality has also been associated with α-diversity metrices.^40^ Similarly, we observed a negative association between MBPS and diet quality; however, this was independent of Shannon index or sodium intake (data not shown).

Moreover, we uncovered differentially prevalent bacteria taxa between participants with different metrics of BP variability. Most notably were the levels of *Alistipes finegoldii*, a newly discovered member of the Bacteroidota (previously known as Bacteroidetes) phylum,^41^ which were higher in the three lowest quartiles of BP variability and MBPS. However, there is contradicting evidence for an association between *Alistipes* spp. in CVD.^42^ Shotgun metagenomic analysis of faecal samples from normotensive and hypertensive patients indicated that two specific *Alistipes* spp. – *Alistipes finegoldii* and *Alistipes indistinctus* – were positively correlated with systolic BP,^43^ with *Alistipes finegoldii* specifically associated with intestinal inflammation.^42,43^ Similarly, greater levels of *Lactobacillus* spp., a low abundance species commonly used in probiotics, were observed in normal night-time dippers and participants in the three lowest quartiles of MBPS. In mouse and human studies, *Lactobacillus* spp. levels were reduced by sodium intake and increased BP via pro-inflammatory pathways including interleukin-17.^44^

Contrastingly, we detected greater levels of several *Clostridium* spp. in participants with the highest quartiles for BP SD, MBPS and extreme dippers, all of which embody a dysregulated circadian rhythm of BP variability. Indeed, an array of *Clostridium* spp. are positively associated with systolic BP.^45^ In mice, when the gut microbial rhythmicity becomes compromised, the host exhibited fluctuations in the ‘normal’ cycle of circadian rhythm as well as increased abundance of *Clostridium* spp.^13^ Another more abundant taxa in those with higher BP variability was *Prevotella* spp., abundantly present in both prehypertensive and hypertensive patients^17^ and identified as early markers in the development of hypertension and CVD.^10^ Moreover, the diurnal activity of the *Prevotella* spp. shows a moderate percentage abundance (between 30-40%) during night-time and a persistent plummet in relative abundance during rising (∼20%).^46^ This suggests that an extreme night-time dip or morning surge phenotype may, in part, be due to greater levels of *Prevotella* spp.

Bacteria in the gastrointestinal tract are metabolically active, and metabolites, specifically SCFAs such as acetate, butyrate and propionate, have been shown to affect host circadian rhythms.^14,47^ Here we identified an inverse association with overall plasma SCFAs, specifically acetate, and MBPS. This may suggest a potential link between acetate metabolism or utilisation and BP, whereby under metabolic stress, a specific liver enzyme becomes activated and generates free acetate into the circulation, which is followed by rapid uptake by peripheral tissue,^48^ and may in part explain lower plasma acetate in participants with higher MBPS. Importantly, we have previously shown that acetate indeed lowers BP in animal models of hypertension.^49,50^ Moreover, SCFA-producing bacterial species were only found in mice not subjected to circadian disruption,^51^ further supporting an association of gut microbial metabolites and circadian dysregulation.

The exact mechanism through which SCFAs are able to elicit alterations in the circadian clock is not yet fully understood.^52^ SCFAs mostly activate signalling via binding to three G-protein coupled receptors, with GPR43 being the most predominant of these receptors. These receptors are highly expressed in immune cells^37^ and activate anti-inflammatory downstream pathways.^53^ Here we identified a negative association between BP SD and night-time dipping and GPR43 in circulating immune cells. This is consistent with our previous findings regarding 24-h systolic BP^20^ and aortic stiffness,^54^ suggesting a blunted response to BP-lowering metabolites.

We acknowledge that there are some limitations to our study, including the sample size, as well as lack of assessment of the presence of sleep disorders, such as obstructive sleep apnoea, which may affect the BP variability profile. Due to the sample size, men and women were analysed together; however, we adjusted some of the analyses for sex, age and BMI. However, our study took advantage of the only multi-site cohort published to date with well-characterised ambulatory BP monitoring that includes data for BP variability (represented as SD), night-time dipping and MBPS in both men and women, all untreated for BP-lowering medication. This is also the only cohort that contains detailed information regarding diet, plasma and faecal SCFAs, and their receptors, which allowed us to explore the interplay between the gut microbiome, diet, their metabolites and receptors in relation to three separate parameters of BP variability. Another limitation is that this is a cross-sectional study, which infers association rather than causation.

### Perspectives

While there was no change in gut microbial diversity in patterns of BP variation over a 24-hour period, we identified significant shifts in bacterial abundances associated with BP variability, MBPS or night-time dipping dippers. To confirm causation, reverse microbiome approaches using germ-free animals^11^ with inoculation of specific taxa identified here, such as *Clostridium* spp. and *Prevotella* spp., would advance our understanding of how specific microbes contribute to BP variability. An important mechanism may be driven by SCFAs, particularly acetate, and its main sensing receptor, GPR43, which were associated with BP variability. Lack of GPR43 may blunt response to BP-lowering metabolites, and may represent a new target for BP therapy in the future.

## Data Availability

Anonymized microbiome data and materials have been made publicly available at the NCBI Sequence Read Archive (SRA) database under access PRJNA722359

https://www.ncbi.nlm.nih.gov/bioproject/PRJNA722359/

## Acknowledgements

We are grateful to Donna Vizi, Vivian Mak and Kaye Carter who assisted with blood collection.

## Novelty and Significance

### 1. What is new?

- This is the first study to assess the gut microbiome, gut microbial metabolites called short-chain fatty acids and their receptors (GPR41/43/109A) in relation to three separate parameters of blood pressure variability (standard deviation, night-time dipping and morning surge).
- We identified specific microbial signatures associated with blood pressure variability in humans.
- The taxa we identified may impact blood pressure variability via mechanisms that involve microbial metabolites, such as the short-chain fatty acid acetate, and their receptor GPR43.

### 2. What is relevant?

- There is limited research into the interplay between the gut microbiome, and more specifically, the metabolites they produce and their metabolite-sensing counterparts in the context of human blood pressure variability.
- Gut bacteria and their metabolites may affect blood pressure variability via systemic mechanisms outside the intestine.
- Targeting these bacteria and associated metabolites may lead to new therapies to reduce blood pressure variability; however, this needs to be tested in clinical trials.

