## Supplementary material for "The Gut Microbiome and their Metabolites in Human Blood Pressure Variability": Online supplemental tables and figures

^1^Hypertension Research Laboratory, School of Biological Sciences, Monash University, Melbourne, Australia; ^2^Department of Gastroenterology, Monash University, Melbourne, Australia; ^3^School of Life and Environmental Sciences, Charles Perkins Centre, University of Sydney, Sydney, Australia; ^4^Preclinical Disease and Prevention, Baker Heart and Diabetes Institute, Melbourne, Australia; ^5^Neuropharmacology Laboratory, Baker Heart and Diabetes Institute, Melbourne, Australia; ^6^Central Clinical School, Faculty of Medicine Nursing and Health Sciences, Monash University, Melbourne, Australia; ^7^Department of Cardiology, Alfred Hospital, Melbourne, Australia; ^8^Department of Pharmacology, Faculty of Medicine Nursing and Health Sciences, Monash University, Melbourne, Australia; ^9^Heart Failure Research Group, Baker Heart and Diabetes Institute, Melbourne, Australia.

**Online Supplementary Tables**

**Table S1.** Correlation analyses of α diversity metrics with all parameters of BP variability.

| **Parameter** | **Observed** | | **Chao1** | | **Shannon** | |
| --- | --- | --- | --- | --- | --- | --- |
|  | Correlation coefficient (r) | *P-*value | Correlation coefficient (r) | *P-*value | Correlation coefficient (r) | *P-*value |
| SD of day fitted data | 0.066 | 0.60 | 0.062 | 0.61 | 0.075 | 0.55 |
| SD of night fitted data | -0.11 | 0.36 | -0.11 | 0.36 | -0.16 | 0.19 |
| SD of total data fitted | 0.021 | 0.86 | 0.018 | 0.88 | -0.058 | 0.64 |
| Night-time dipping | -0.056 | 0.65 | -0.030 | 0.81 | -0.17 | 0.17 |
| Sleep-through MBPS | -0.021 | 0.87 | -0.004 | 0.98 | -0.12 | 0.32 |
| Prewaking MBPS | -0.14 | 0.27 | -0.12 | 0.34 | -0.24 | **0.046** |

Legend: standard deviation, SD; morning blood pressure surge, MBPS. In bold, P<0.05 which was considered statistically significant.

**Table S2.** **Taxonomic features associated with blood pressure variability**, measured as standard deviation (SD).

| **Taxonomic feature** | **log2FC** | ***P*-value** | **FDR** | **Variable** |
| --- | --- | --- | --- | --- |
| EnterobacterspUCDUG_FMILLET | 4.46 | 3.37E-20 | 9.40E-18 | SD total |
| **unculturedClostridiabacterium_5** | 3.79 | 8.29E-15 | 2.3E-12 | SD day |
| **unculturedThermoanaerobacteralesbacterium** | 3.35 | 1.66E-13 | 2.30E-11 | SD total |
| **Enterobacteriaceae** | 3.85 | 2.29E-10 | 3.2E-08 | SD day |
| **unculturedClostridiabacterium_5** | 3.12 | 2.18E-09 | 2.00E-07 | SD total |
| **ClostridialesvadinBB60group** | 3.55 | 4.85E-08 | 3.40E-06 | SD total |
| Parasutterellasecunda | 2.97 | 3.84E-08 | 3.6E-06 | SD day |
| **Enterobacteriaceae** | 3.27 | 2.06E-07 | 1.10E-05 | SD total |
| **Paraprevotella** | -4.90 | 3.01E-06 | 0.00014 | SD total |
| metagenome_1 | -5.24 | 6.23E-06 | 0.00025 | SD total |
| **ClostridialesvadinBB60group** | 3.07 | 5.11E-06 | 0.00029 | SD day |
| ParabacteroidesspYL27 | -5.63 | 4.46E-06 | 0.00029 | SD day |
| metagenome_22 | 2.10 | 2.92E-06 | 0.0008 | SD night |
| **gutmetagenome_32** | 2.70 | 2.34E-05 | 0.0011 | SD day |
| **Azospirillumsp47_25** | -3.62 | 3.59E-05 | 0.0013 | SD total |
| **unculturedrumenbacterium5C0d12** | 2.93 | 3.54E-05 | 0.0014 | SD day |
| unculturedClostridiumsp_8 | 2.22 | 4.60E-05 | 0.0016 | SD day |
| **uncultured_1** | 1.77 | 6.73E-05 | 0.0019 | SD day |
| **unculturedRuminococcussp** | 2.69 | 6.01E-05 | 0.0019 | SD day |
| **unculturedAcetivibriosp** | -4.62 | 6.64E-05 | 0.0021 | SD total |
| gutmetagenome_12 | 2.27 | 3.17E-05 | 0.0022 | SD night |
| **gutmetagenome_32** | 2.69 | 2.33E-05 | 0.0022 | SD night |
| MollicutesRF39 | 2.59 | 2.27E-05 | 0.0022 | SD night |
| unculturedFirmicutesbacterium_1 | -2.61 | 7.85E-05 | 0.0022 | SD total |
| RikenellaceaeRC9gutgroup | -5.60 | 9.13E-05 | 0.0023 | SD day |
| Anaerofilum | -2.68 | 0.000111 | 0.0026 | SD day |
| **unculturedrumenbacterium5C0d12** | -4.15 | 8.56E-05 | 0.0048 | SD night |
| Collinsella | 1.32 | 0.000242 | 0.0052 | SD day |
| **metagenome_21** | 2.40 | 0.000257 | 0.0056 | SD total |
| unculturedClostridiumsp_6 | 1.53 | 0.000262 | 0.0056 | SD total |
| **unculturedrumenbacterium5C0d12** | -3.84 | 0.00026 | 0.0056 | SD total |
| **Lactobacillus** | -4.26 | 0.000366 | 0.0073 | SD total |
| **uncultured_1** | 1.57 | 0.000392 | 0.0073 | SD total |
| LachnospiraceaeUCG001 | -2.55 | 0.000378 | 0.0075 | SD day |
| **metagenome_21** | 2.46 | 0.000171 | 0.008 | SD night |
| RuminococcusspID1 | -2.42 | 0.000535 | 0.0099 | SD day |
| Phascolarctobacterium | -2.89 | 0.00063 | 0.01 | SD total |
| unculturedrumenbacterium_3 | -2.64 | 0.000613 | 0.01 | SD total |
| BurkholderiaspK4410MGS135 | -3.15 | 0.000721 | 0.010 | SD day |
| **CandidatusGastranaerophilalesbacteriumZag_111** | -3.00 | 0.000628 | 0.010 | SD day |
| gutmetagenome_30 | 2.61 | 0.000629 | 0.010 | SD day |
| metagenome_18 | -2.61 | 0.000678 | 0.010 | SD day |
| Pasteurellaceae | -2.36 | 0.000719 | 0.010 | SD day |
| unculturedClostridiumsp_4 | -2.21 | 0.000883 | 0.012 | SD day |
| **Paraprevotella** | -3.34 | 0.001149 | 0.015 | SD day |
| **unculturedorganism_13** | 1.34 | 0.001009 | 0.016 | SD total |
| unculturedorganism_8 | 1.37 | 0.001345 | 0.016 | SD day |
| **unculturedThermoanaerobacteralesbacterium** | 1.75 | 0.001367 | 0.016 | SD day |
| **Clostridiales** | 1.86 | 0.001133 | 0.017 | SD total |
| **Alistipesfinegoldii** | -3.08 | 0.000444 | 0.018 | SD night |
| gutmetagenome_41 | 2.08 | 0.001274 | 0.018 | SD total |
| gutmetagenome_17 | 2.31 | 0.001956 | 0.022 | SD day |
| gutmetagenome_2 | -3.61 | 0.000778 | 0.022 | SD night |
| metagenome_19 | -3.19 | 0.00066 | 0.022 | SD night |
| Ruminococcus1 | 1.89 | 0.00074 | 0.022 | SD night |
| BacteroidesspSmarlab3302398 | 1.82 | 0.000932 | 0.023 | SD night |
| **unculturedorganism_13** | 1.35 | 0.000983 | 0.023 | SD night |
| unculturedorganism_12 | 1.25 | 0.002258 | 0.024 | SD day |
| **Lactobacillus** | -3.68 | 0.002433 | 0.025 | SD day |
| **Lactobacillus** | -3.82 | 0.001204 | 0.025 | SD night |
| Turicibacter | 1.36 | 0.001349 | 0.025 | SD night |
| **unculturedrumenbacterium4C0d9** | 1.39 | 0.001272 | 0.025 | SD night |
| **unculturedRuminococcussp** | -3.07 | 0.001443 | 0.025 | SD night |
| gutmetagenome_25 | -2.08 | 0.001961 | 0.026 | SD total |
| **Azospirillumsp47_25** | -2.57 | 0.003155 | 0.031 | SD day |
| Clostridialesbacterium607e | 1.35 | 0.001963 | 0.031 | SD night |
| Clostridialesbacteriumcanineoraltaxon162 | -3.84 | 0.002 | 0.031 | SD night |
| unculturedeubacteriumWCHB154 | 1.45 | 0.002469 | 0.031 | SD total |
| unculturedorganism_19 | 1.64 | 0.00318 | 0.031 | SD day |
| **Alistipesfinegoldii** | 1.90 | 0.003423 | 0.032 | SD day |
| **unculturedAcetivibriosp** | -3.40 | 0.002214 | 0.033 | SD night |
| gutmetagenome_26 | -2.56 | 0.003891 | 0.035 | SD day |
| unculturedprokaryote_2 | 1.66 | 0.004094 | 0.036 | SD day |
| Faecalibacteriumprausnitzii | -1.80 | 0.00441 | 0.037 | SD day |
| RuminococcaceaeUCG009 | 1.27 | 0.004527 | 0.037 | SD day |
| **Clostridiales** | 1.73 | 0.002772 | 0.039 | SD night |
| gutmetagenome_31 | -3.42 | 0.004854 | 0.039 | SD day |
| BacteroidespectinophilusATCC43243 | 1.62 | 0.003838 | 0.047 | SD total |
| anaerobicdigestermetagenome | -1.52 | 0.004575 | 0.049 | SD total |
| **CandidatusGastranaerophilalesbacteriumZag_111** | -2.38 | 0.00448 | 0.049 | SD total |
| Erysipelatoclostridium | 1.44 | 0.004485 | 0.049 | SD total |
| unculturedBarnesiellasp | 1.94 | 0.004983 | 0.049 | SD total |
| unculturedorganism_4 | 1.75 | 0.005063 | 0.049 | SD total |
| **unculturedrumenbacterium4C0d9** | 1.23 | 0.004927 | 0.049 | SD total |

Legend: FDR, false discovery rate; log2FC, log2 fold change. Taxa in bold are present in association with more than one variable.

**Table S3.** **Taxonomic features associated with all three night-time dipping profiles (extreme dippers, dippers and non-dippers)**.

| **Taxonomic feature** | **log2FC** | ***P*-value** | **FDR** |
| --- | --- | --- | --- |
| ClostridialesvadinBB60group | -5.58 | 4.45E-14 | 1.2E-11 |
| unculturedorganism_19 | -4.16 | 3.89E-12 | 5.5E-10 |
| unculturedFlavonifractorsp_1 | -2.91 | 6.08E-07 | 5.7E-05 |
| PrevotellaceaeNK3B31group | -3.69 | 0.00030036 | 0.017 |
| gutmetagenome_18 | 5.39 | 0.00031655 | 0.017 |
| metagenome_9 | 4.32 | 0.00036967 | 0.017 |
| Gaboniamassiliensis | -2.23 | 0.00050734 | 0.020 |
| unculturedRuminococcussp | -2.73 | 0.00090698 | 0.030 |
| Lactobacillus | 6.61 | 0.0009495 | 0.030 |
| unculturedorganism_5 | 3.58 | 0.001092 | 0.031 |

Legend: FDR, false discovery rate; log2FC, log2 fold change.

**Table S4. Taxonomic features associated with dippers vs. extreme dippers.**

| **Taxonomic feature** | **log2FC** | ***P*-value** | **FDR** |
| --- | --- | --- | --- |
| ClostridialesvadinBB60group | 5.38 | 2.35E-13 | 6.81E-11 |
| unculturedorganism_18 | 3.69 | 3.47E-12 | 5.03E-10 |
| unculturedFlavonifractorsp_1 | 2.68 | 6.50E-06 | 0.00063 |
| PrevotellaceaeNK3B31group | 3.57 | 0.0002696 | 0.018 |
| metagenome_8 | -4.47 | 0.0003068 | 0.018 |
| gutmetagenome_17 | -5.36 | 0.0003891 | 0.019 |
| metagenome_16 | -3.58 | 0.001005 | 0.041 |
| MegamonasfuniformisYIT11815 | 3.23 | 0.001253 | 0.041 |
| Bacteroidesmediterraneensis | 3.41 | 0.001374 | 0.041 |
| unculturedRuminococcussp | 2.80 | 0.001423 | 0.041 |
| unculturedorganism_5 | -3.43 | 0.001566 | 0.041 |
| Gaboniamassiliensis | 2.09 | 0.001794 | 0.041 |
| Gastranaerophilales | 2.34 | 0.001830 | 0.041 |

Legend: FDR, false discovery rate; log2FC, log2 fold change.

**Table S5. Taxonomic features associated with dippers vs. non-dippers.**

| **Taxonomic feature** | **log2FC** | ***P*-value** | **FDR** |
| --- | --- | --- | --- |
| ParabacteroidesspYL27 | 3.75 | 7.94E-07 | 0.00010 |
| unculturedorganism_19 | 2.43 | 1.31E-06 | 0.00010 |
| Lactobacillus | -5.89 | 1.47E-06 | 0.00010 |
| unidentifiedrumenbacteriumRFN33 | -4.85 | 1.54E-06 | 0.00010 |
| unculturedClostridiumsp_4 | 2.08 | 1.02E-05 | 0.00055 |
| Clostridiales | 2.49 | 1.39E-05 | 0.00063 |
| metagenome_21 | 2.75 | 4.27E-05 | 0.0016 |
| gutmetagenome_25 | 2.64 | 5.01E-05 | 0.0017 |
| unculturedrumenbacterium5C0d12 | 2.90 | 9.22E-05 | 0.0028 |
| Sanguibacteroides | 1.66 | 0.0001115 | 0.0030 |
| unculturedorganism_11 | 1.57 | 0.0003168 | 0.0078 |
| Sutterella | -2.65 | 0.0006485 | 0.014 |
| gutmetagenome_22 | -2.30 | 0.0006692 | 0.014 |
| Phascolarctobacterium | -2.75 | 0.0009095 | 0.018 |
| PrevotellaceaeNK3B31group | 2.84 | 0.001026 | 0.018 |
| rumenbacteriumNK4A214 | 1.92 | 0.001275 | 0.022 |
| Lachnoclostridium | -2.48 | 0.001371 | 0.022 |
| unculturedRuminococcussp | -2.64 | 0.001511 | 0.023 |
| metagenome_1 | -3.33 | 0.001649 | 0.023 |
| uncultured_1 | -1.79 | 0.002932 | 0.039 |
| metagenome_19 | -2.75 | 0.003153 | 0.039 |
| gutmetagenome_31 | 2.04 | 0.003178 | 0.039 |
| BacteroidesspHPS0048 | -2.42 | 0.003727 | 0.044 |
| Ruminiclostridium6 | 1.95 | 0.004078 | 0.046 |

Legend: FDR, false discovery rate; log2FC, log2 fold change.

**Table S6.** **Taxonomic features associated with morning blood pressure surge**.

| **Taxonomic feature** | **log2FC** | ***P*-value** | **FDR** | **Variable** |
| --- | --- | --- | --- | --- |
| **Lactobacillus** | 5.73 | 5.62E-16 | 1.56E-13 | Prewaking |
| **ClostridialesvadinBB60group** | 4.07 | 1.72E-10 | 2.39E-08 | Prewaking |
| **Roseburia** | 3.89 | 1.02E-08 | 2.80E-06 | Sleep-through |
| **gutmetagenome_32** | 3.37 | 4.07E-08 | 5.70E-06 | Sleep-through |
| ParabacteroidesspYL27 | -5.89 | 7.21E-07 | 6.70E-05 | Sleep-through |
| unculturedorganism_1 | -6.34 | 7.67E-07 | 7.11E-05 | Prewaking |
| **ClostridialesvadinBB60group** | 3.23 | 1.48E-06 | 0.0001 | Sleep-through |
| **unculturedRuminococcussp** | -4.71 | 3.36E-06 | 0.00023 | Prewaking |
| **unculturedRuminococcussp** | -4.60 | 4.49E-06 | 0.00025 | Sleep-through |
| metagenome_21 | 2.82 | 1.12E-05 | 0.00062 | Prewaking |
| **gutmetagenome_2** | -4.54 | 4.20E-05 | 0.0017 | Sleep-through |
| **Lactobacillus** | -5.10 | 3.58E-05 | 0.0017 | Sleep-through |
| bacteriumNLAEzlH46 | 2.64 | 5.18E-05 | 0.0018 | Prewaking |
| unculturedbacterium_2 | -3.27 | 5.01E-05 | 0.0018 | Prewaking |
| unculturedrumenbacterium4C0d9 | 1.72 | 4.49E-05 | 0.0018 | Prewaking |
| unculturedrumenbacterium5C0d12 | -4.23 | 6.41E-05 | 0.0022 | Sleep-through |
| Enterobacteriaceae | -3.62 | 0.00015596 | 0.0048 | Sleep-through |
| CandidatusGastranaerophilalesbacteriumZag_111 | -3.21 | 0.00022315 | 0.0062 | Prewaking |
| metagenome_7 | 1.76 | 0.00021613 | 0.0062 | Prewaking |
| **gutmetagenome_32** | 2.38 | 0.00025987 | 0.0063 | Prewaking |
| unculturedbacteriumadhufec58 | -3.55 | 0.00027294 | 0.0063 | Prewaking |
| Lachnoclostridium | 2.10 | 0.00033242 | 0.0066 | Prewaking |
| Prevotella9 | 2.85 | 0.00031055 | 0.0066 | Prewaking |
| Clostridiales | 2.04 | 0.00036381 | 0.0067 | Prewaking |
| BacteroidesplebeiusDSM17135 | -4.78 | 0.00040982 | 0.0071 | Prewaking |
| Dialister | -3.48 | 0.00031227 | 0.0087 | Sleep-through |
| unculturedAcetivibriosp | -4.06 | 0.0003525 | 0.0089 | Sleep-through |
| rumenbacteriumNK4A214 | -2.66 | 0.00039326 | 0.0091 | Sleep-through |
| BurkholderiaspK4410MGS135 | 2.24 | 0.00056086 | 0.0092 | Prewaking |
| Clostridialesbacteriumcanineoraltaxon162 | -4.45 | 0.00046932 | 0.01 | Sleep-through |
| Massiliprevotellamassiliensis | -3.78 | 0.00078177 | 0.011 | Prewaking |
| metagenome_15 | 2.00 | 0.00076999 | 0.011 | Prewaking |
| unculturedClostridiumsp_4 | -2.16 | 0.00070066 | 0.011 | Prewaking |
| MollicutesRF39 | 2.07 | 0.00096382 | 0.013 | Prewaking |
| bacteriummpnisolategroup19 | -2.16 | 0.0013723 | 0.016 | Prewaking |
| unculturedClostridialesFamilyXIIIbacterium | 1.37 | 0.0012395 | 0.016 | Prewaking |
| unculturedorganism_3 | 2.42 | 0.0013211 | 0.016 | Prewaking |
| unculturedorganism_4 | -2.58 | 0.0019682 | 0.022 | Prewaking |
| **gutmetagenome_2** | -3.25 | 0.0022102 | 0.024 | Prewaking |
| LachnospiraceaeUCG001 | -2.08 | 0.0025475 | 0.026 | Prewaking |
| **EnterobacterspUCDUG_FMILLET** | -2.48 | 0.002719 | 0.027 | Prewaking |
| Alistipesfinegoldii | -2.76 | 0.0015652 | 0.029 | Sleep-through |
| **EnterobacterspUCDUG_FMILLET** | -2.64 | 0.0015768 | 0.029 | Sleep-through |
| metagenome_4 | -2.30 | 0.0017108 | 0.03 | Sleep-through |
| metagenome_1 | -3.10 | 0.0039591 | 0.038 | Prewaking |
| Agathobacter | 1.75 | 0.0024266 | 0.039 | Sleep-through |
| AlistipesspN15MGS157 | -2.49 | 0.0042455 | 0.039 | Prewaking |
| unidentified_6 | -2.38 | 0.0025123 | 0.039 | Sleep-through |
| BacteroidespectinophilusATCC43243 | 1.66 | 0.0027707 | 0.041 | Sleep-through |
| Bacteroidales | 2.00 | 0.0030793 | 0.043 | Sleep-through |
| Erysipelatoclostridium | -1.82 | 0.0052909 | 0.045 | Prewaking |
| PrevotellaceaeNK3B31group | -3.42 | 0.0053926 | 0.045 | Prewaking |
| **Roseburia** | -2.85 | 0.0052971 | 0.045 | Prewaking |
| unculturedClostridiumsp_8 | 1.57 | 0.005652 | 0.046 | Prewaking |

Legend: FDR, false discovery rate; log2FC, log2 fold change. Taxa in bold are present in association with more than one variable.

**Table S7.** **Faecal and plasma short-chain fatty acids (SCFAs) levels correlated with all parameters of BP variability.**

| **Parameter** | **SD of day fitted data** | | **SD of night fitted data** | | **SD of total fitted data** | | **Night-time Dipping** | | **Sleep-through MBPS** | | **Prewaking MBPS** | |
| --- | --- | --- | --- | --- | --- | --- | --- | --- | --- | --- | --- | --- |
| *SCFAs* | Correlation coefficient (r) | *P*-value | Correlation coefficient (r) | *P*-value | Correlation coefficient (r) | *P*-value | Correlation coefficient (r) | *P*-value | Correlation coefficient (r) | *P*-value | Correlation coefficient (r) | *P*-value |
| *Faecal SCFAs* | | | | | | | | | | | | |
| Acetic acid | 0.062 | 0.64 | 0.21 | 0.10 | 0.19 | 0.14 | 0.039 | 0.76 | 0.11 | 0.40 | -0.006 | 0.97 |
| Propionic acid | -0.042 | 0.75 | 0.090 | 0.49 | 0.067 | 0.61 | 0.075 | 0.56 | 0.085 | 0.52 | 0.084 | 0.52 |
| Butyric acid | -0.005 | 0.97 | 0.15 | 0.25 | 0.094 | 0.47 | 0.015 | 0.91 | 0.021 | 0.88 | 0.006 | 0.96 |
| Total SCFAs | 0.008 | 0.95 | 0.15 | 0.24 | 0.14 | 0.27 | 0.044 | 0.73 | 0.11 | 0.40 | 0.020 | 0.88 |
| *Plasma SCFAs* | | | | | | | | | | | | |
| Acetate | 0.085 | 0.52 | -0.13 | 0.34 | 0.16 | 0.23 | 0.021 | 0.88 | -0.21 | 0.11 | -0.31 | **0.017** |
| Butyrate | 0.092 | 0.49 | 0.11 | 0.42 | 0.12 | 0.38 | -0.041 | 0.76 | 0.051 | 0.70 | -0.076 | 0.57 |
| Propionate | -0.044 | 0.74 | -0.16 | 0.24 | -0.033 | 0.80 | -0.001 | 1.0 | -0.15 | 0.27 | -0.12 | 0.36 |
| Total SCFAs | 0.080 | 0.55 | -0.12 | 0.36 | 0.16 | 0.24 | 0.020 | 0.88 | -0.21 | 0.12 | -0.31 | **0.020** |

In bold, *P*<0.05 considered statistically significant also adjusted for age, sex and BMI in regression analyses.

**Table S8.** **mRNA expression of G-protein coupled receptors *FFAR2, FFAR3* and *HCAR2* correlated with all parameters of BP variability.**

| **Parameter** | ***FFAR3* (GPR41)** | | ***FFAR2* (GPR43)** | | ***HCAR2* (GPR109A)** | |
| --- | --- | --- | --- | --- | --- | --- |
|  | Correlation coefficient (r) | *P*-value | Correlation coefficient (r) | *P*-value | Correlation coefficient (r) | *P*-value |
| SD of day fitted data | -0.19 | 0.22 | -0.31 | **0.039** | 0.085 | 0.59 |
| SD of night fitted data | -0.072 | 0.64 | -0.36 | **0.016** | 0.022 | 0.89 |
| SD of total data fitted | -0.042 | 0.78 | -0.32 | **0.035** | -0.005 | 0.98 |
| Night-time dipping | 0.22 | 0.14 | 0.38 | **0.0087** | -0.12 | 0.42 |
| Sleep-through MBPS | 0.055 | 0.72 | 0.14 | 0.37 | -0.035 | 0.83 |
| Prewaking MBPS | 0.067 | 0.67 | 0.087 | 0.57 | -0.17 | 0.27 |

Legend: standard deviation, SD; morning blood pressure surge, MBPS. In bold, *P*<0.05 considered statistically significant.

**Online Supplementary Figures**

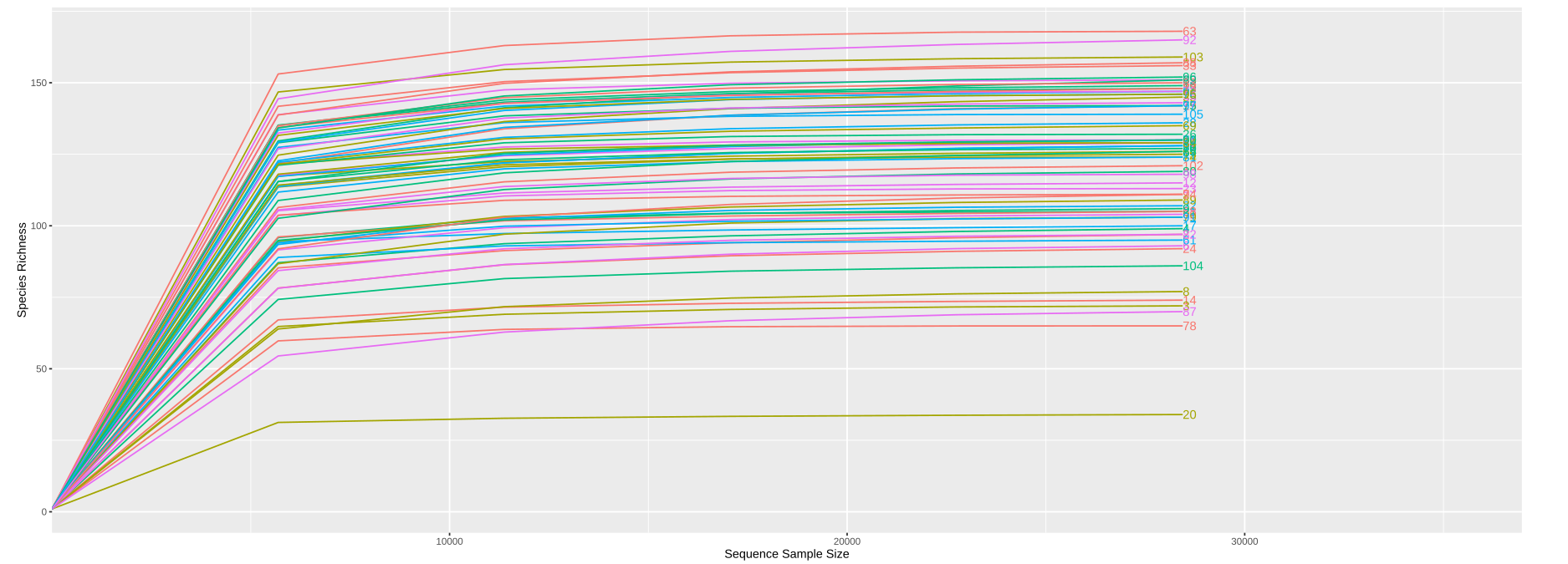
**Figure S1. Rarefaction curve of all samples.** One sample was excluded from the data analysis due to low number of sequencing reads (<10,000).

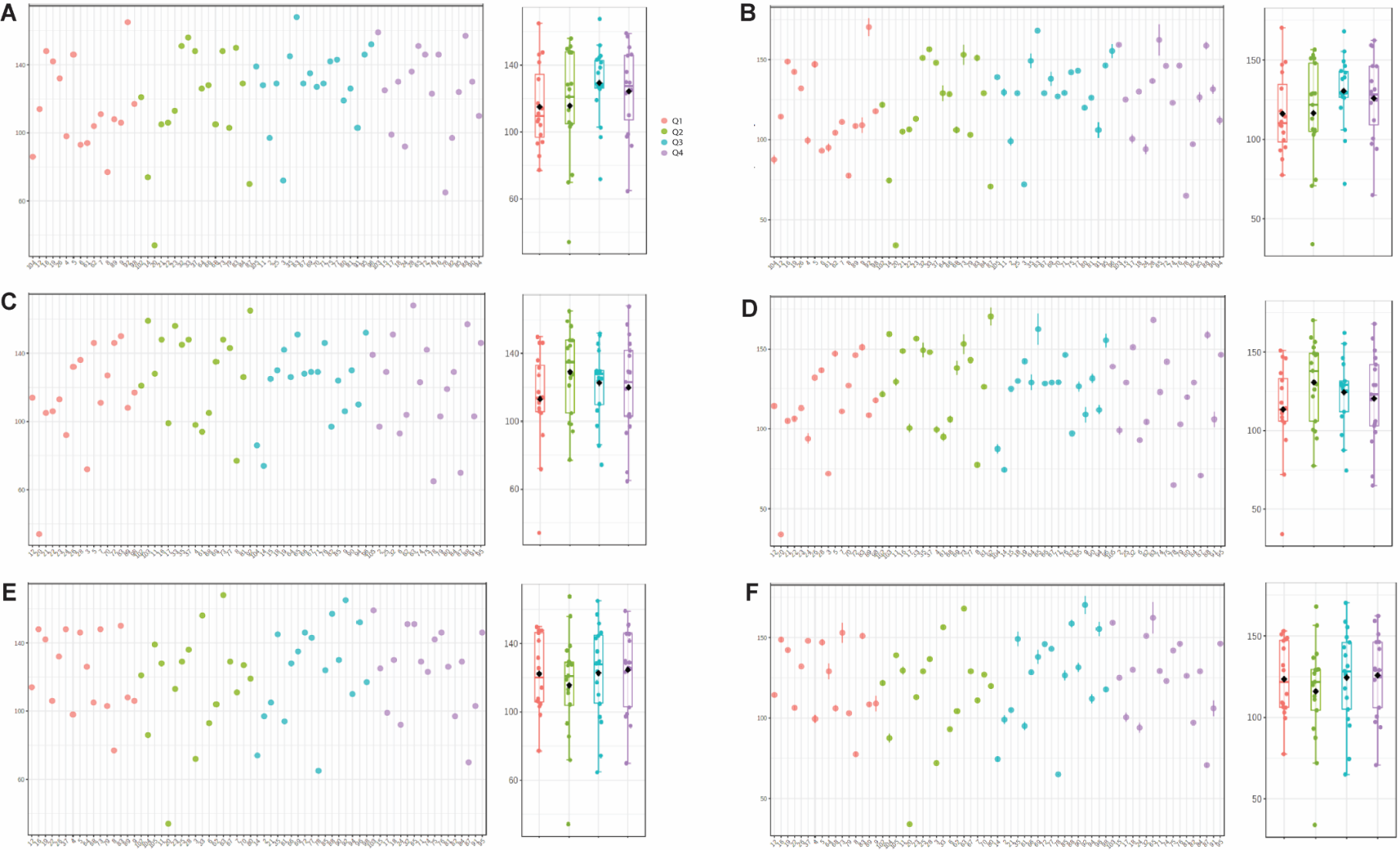

**Figure S2. α-diversity principal coordinate analysis plots for blood pressure standard deviation (SD).** α-diversity profiling showing observed OTUs and Chao1 index for A, B) SD day, C, D) SD night and E, F) total respectively. Box plot data presented as median and inter-quartile range (IQR). Red=Q1, Green=Q2, Blue=Q3 and Purple=Q4. All *P*>0.05.

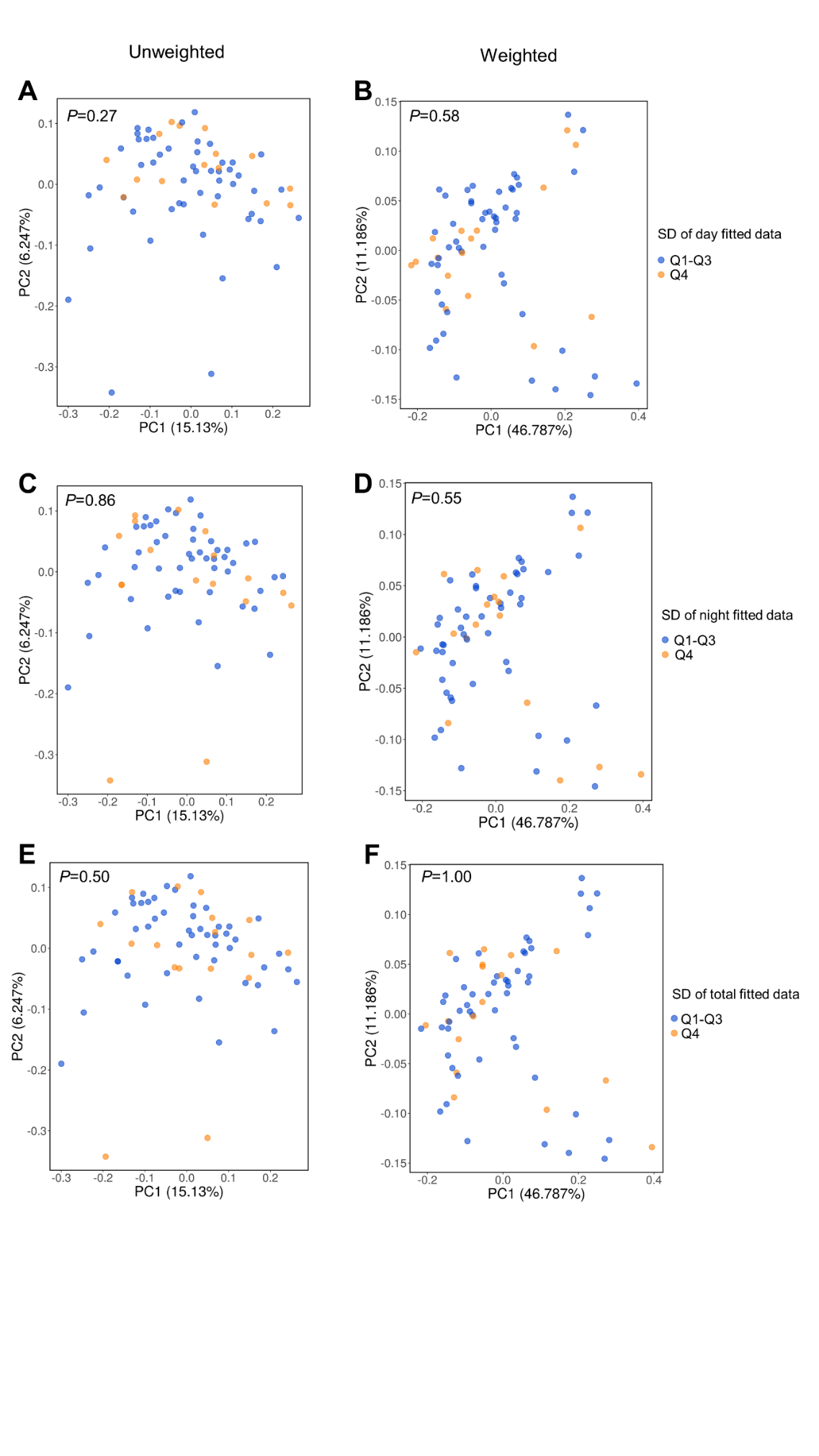

**Figure S3. β-diversity principal coordinate analysis plots for blood pressure standard deviation (SD)**. β-diversity principal coordinate analysis plots of unweighted (i.e., microbial diversity based on presence/absence) and weighted (i.e., microbial diversity based on abundance) UniFrac analyses of A & B) SD of day data fitted, C & D) SD of night data fitted and E & F) SD of total data fitted of Q1-Q3 versus Q4.

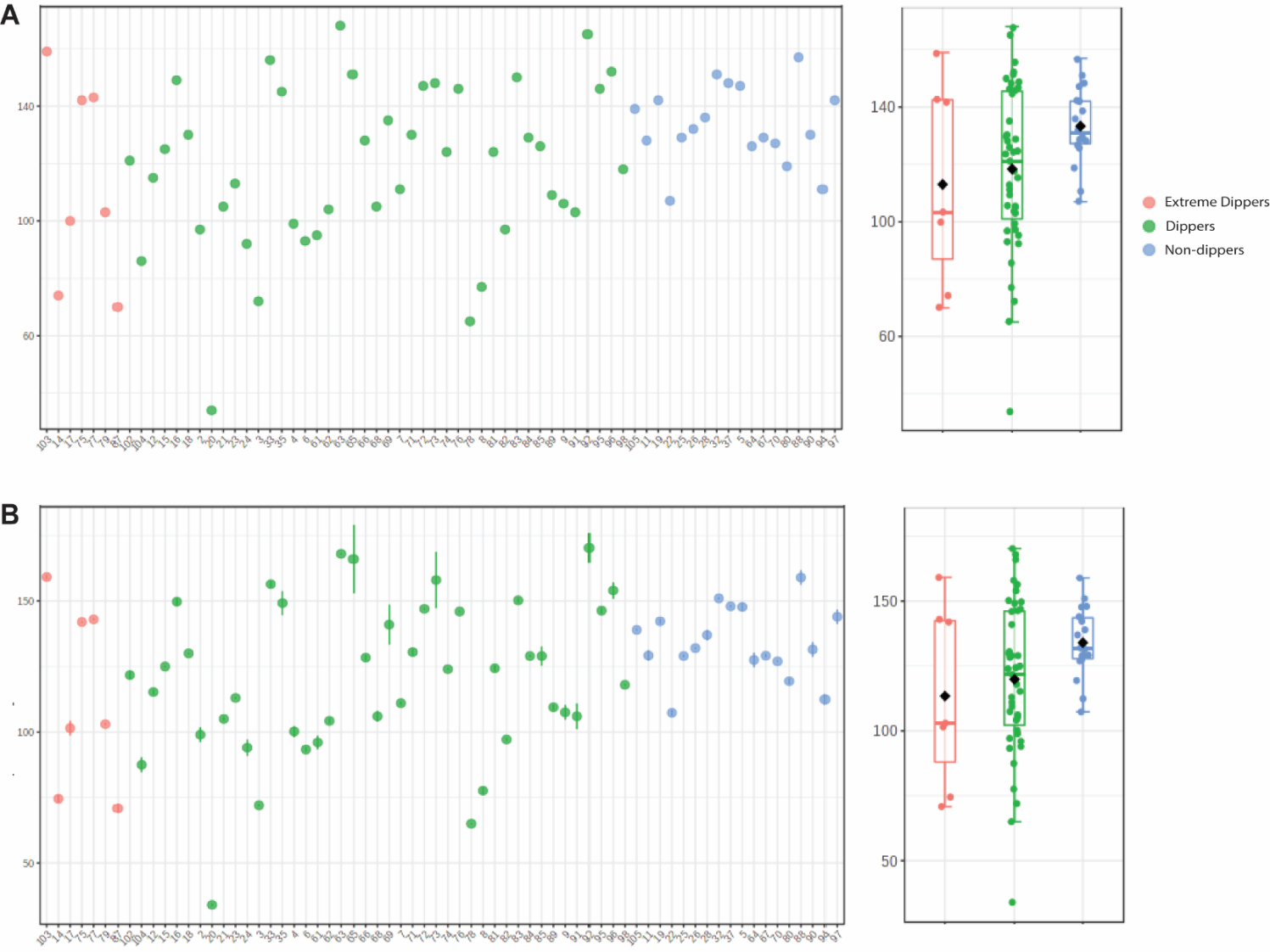

**Figure S4. α-diversity principal coordinate analysis plots night-time dipping**. α-diversity profiling showing A) observed OTUs and B) Chao1 index for night-time dipping. Box plot data presented as median and inter-quartile range (IQR). All *P*>0.05.

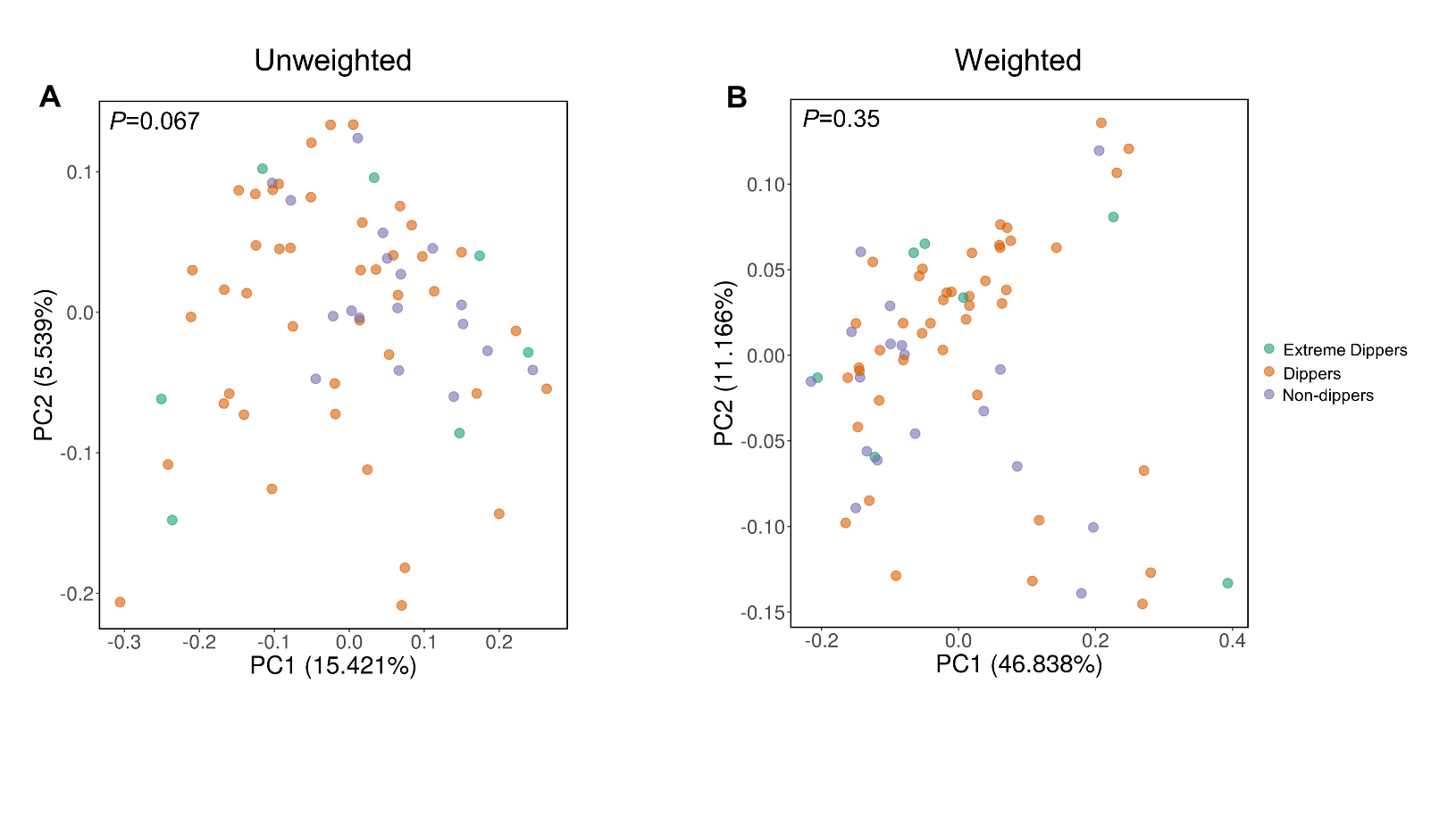
**Figure S5. β-diversity principal coordinate analysis plots for night-time dipping**. β-diversity principal coordinate analysis plots of A) unweighted and B) weighted UniFrac analyses of night-time dipping classifications.

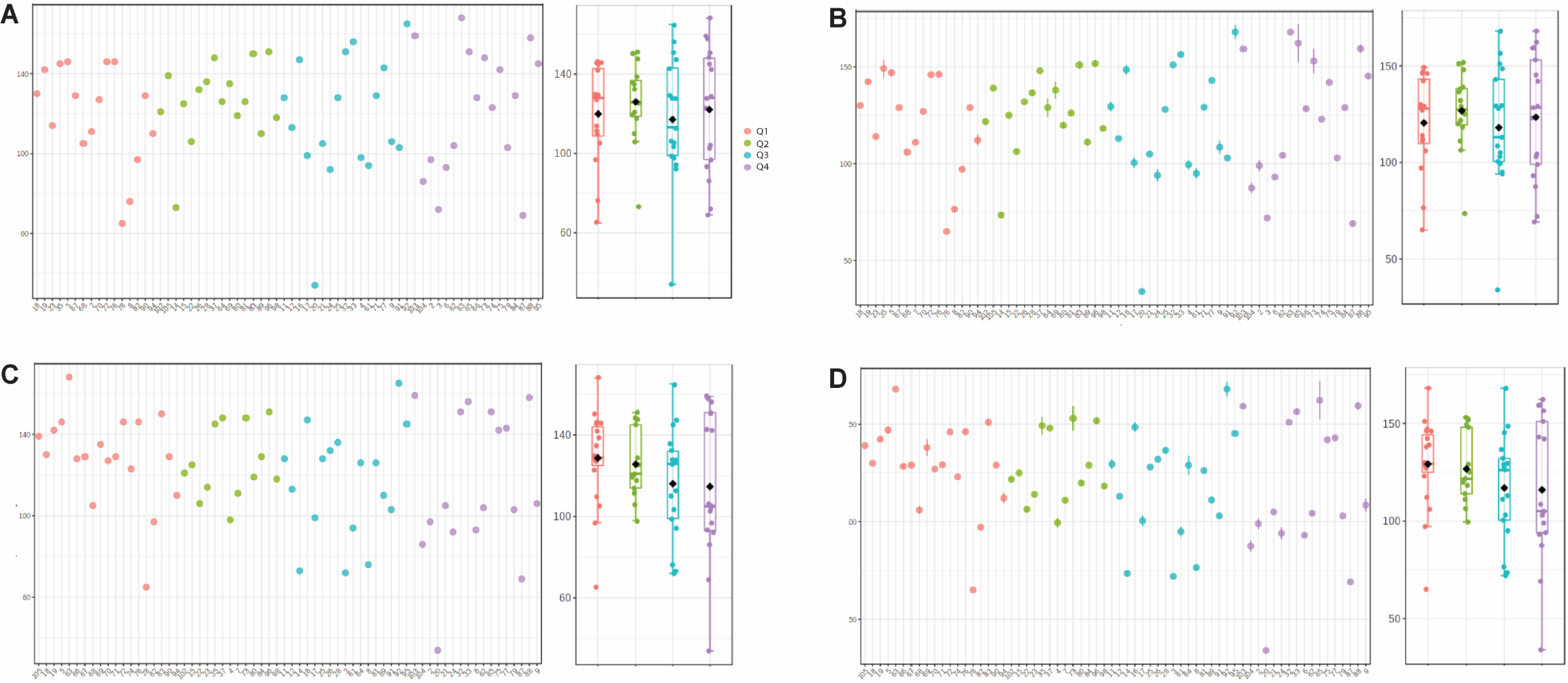

**Figure S6. α-diversity principal coordinate analysis plots for morning blood pressure surge (MBPS)**. α-diversity profiling showing observed OTUs and Chao1 index for A, B) sleep-through and C, D) prewaking MBPS respectively. Box plot data presented as median and inter-quartile range (IQR). Red=Q1, Green=Q2, Blue=Q3 and Purple=Q4. All *P*>0.05.

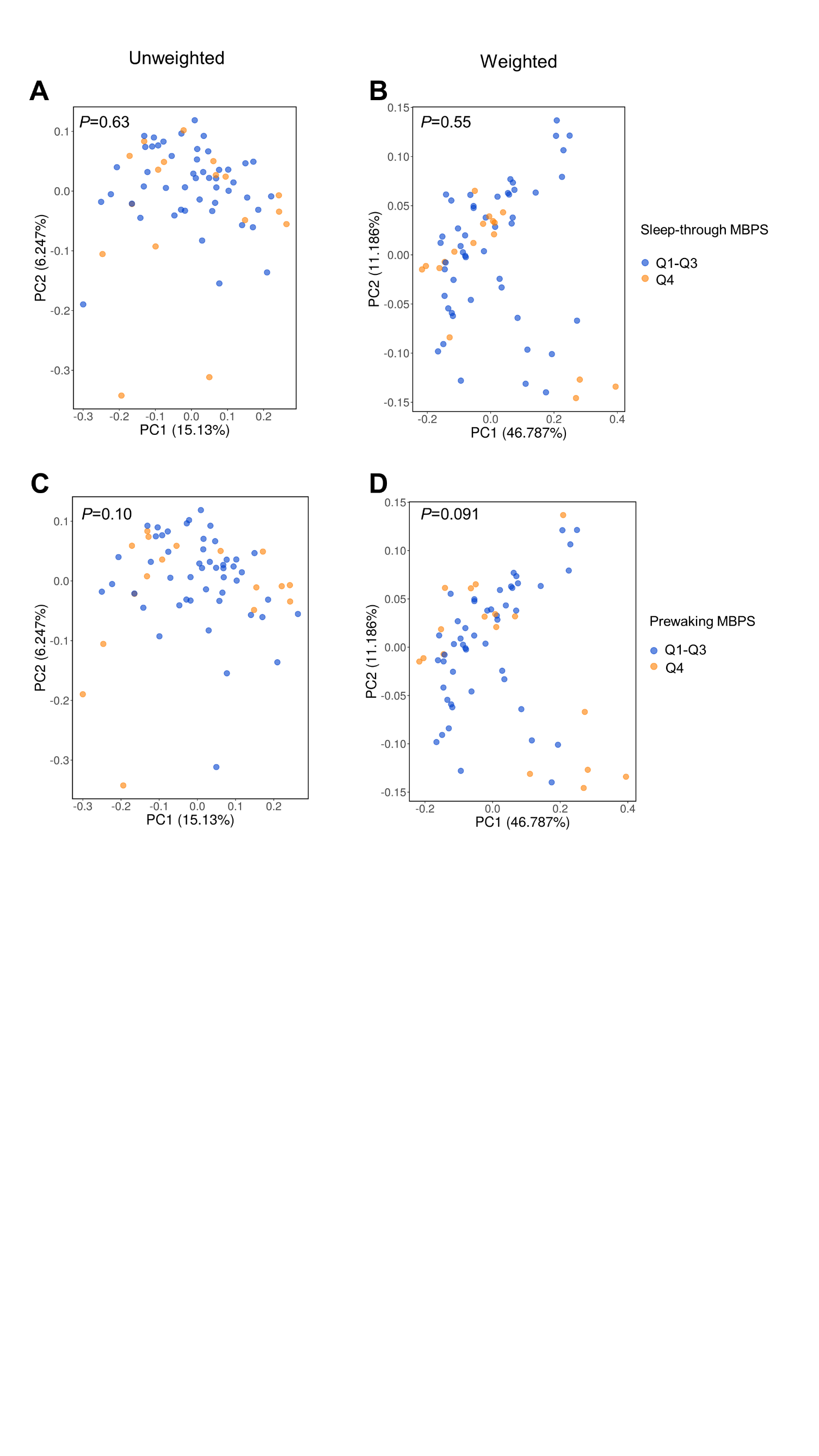

**Figure S7. β-diversity principal coordinate analysis plots for morning blood pressure surge (MBPS).** β-diversity principal coordinate analysis plots of A) unweighted and B) weighted UniFrac analyses of sleep-through MBPS and C) unweighted and B) weighted UniFrac analyses of prewaking MBPS of Q1-Q3 versus Q4.

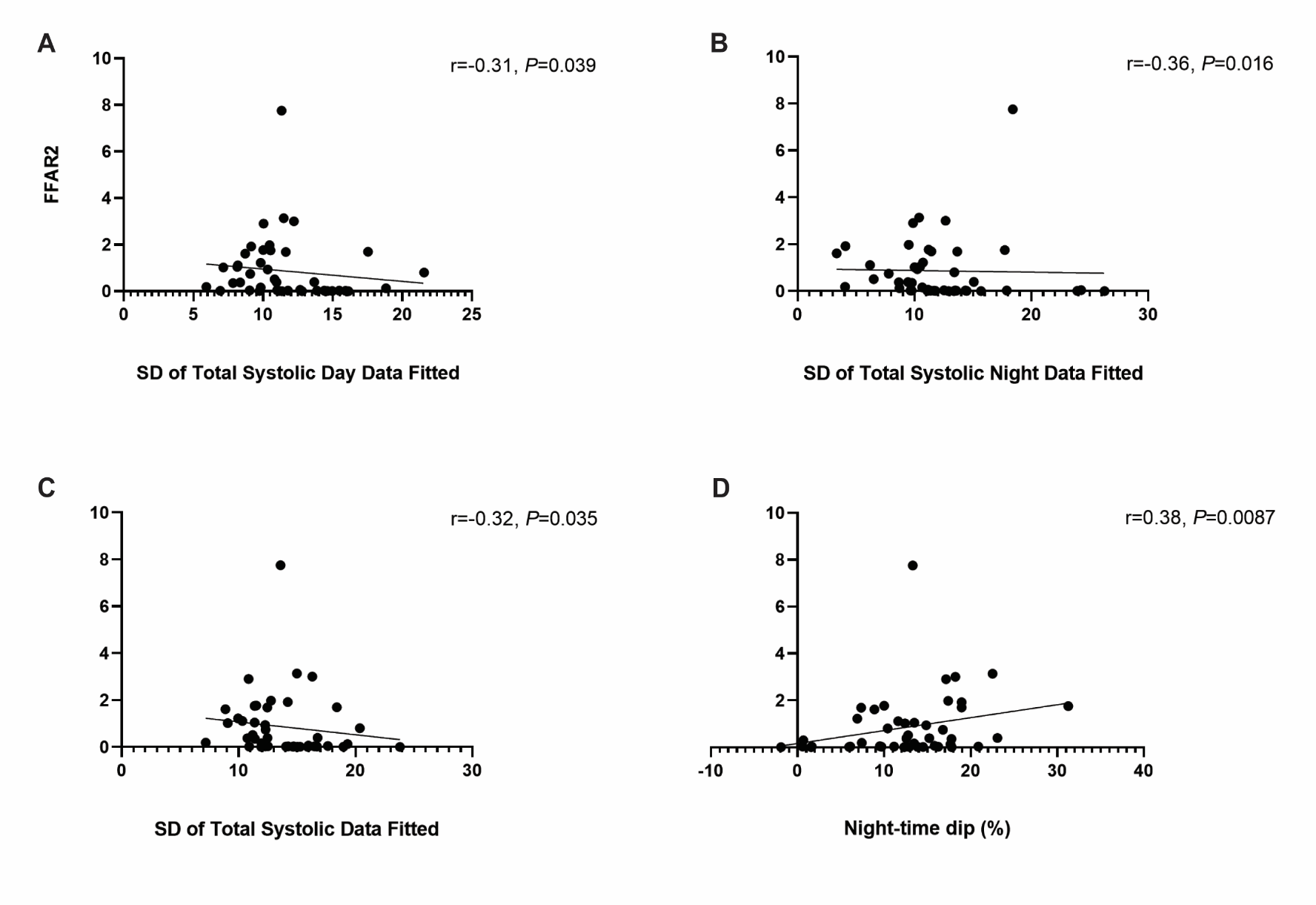

**Figure S8. Correlations between FFAR2 and blood pressure variability**. Spearman correlation analyses between FFAR2 and A) SD day, B) SD night, C) SD total and D) night-time dipping (%).

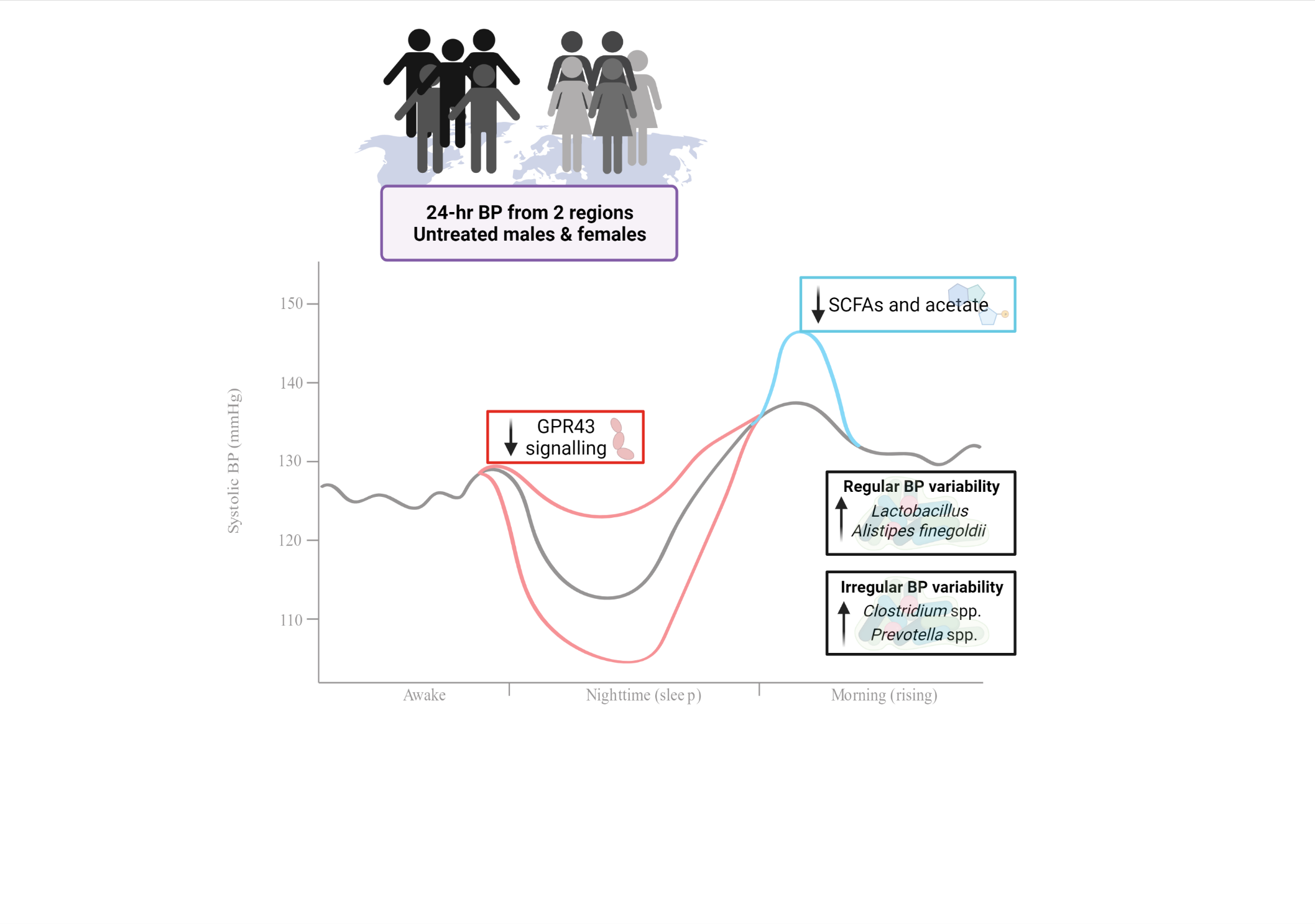
**Figure S9. Summary figure.**
